# Fibrosing interstitial lung disease in childhood: prevalence and disease trajectories

**DOI:** 10.1101/2025.10.24.25338616

**Authors:** Matthias Griese, Simone Reu-Hofer, Julia Ley-Zaporozhan, Birgit Kammer, Ingrid Krüger-Stollfuß, Srdjan Micic, Julia Carlens, Pia Maier, Julia Rodler, Katharina Mauss-Schwarzer, Nguyen-Binh Tran, Christina Katharina Rapp, Florian Gothe, Honorata Marczak, Joanna Lange, Katarzyna Krenke, Astrid Madsen Ring, Frederik Buchvald, Florian Stehling, Pera Silvija Jerkic, Jordis Trischler, Marijke Proesmans, Tugba Sismanlar, Ayse Aslan, Nagehan Emiralioğlu, Nural Kiper, Susanne Hämmerling, Ayşe Kılınç, Freerk Prenzel, Anna Wiemers, Antonio Moreno-Galdo, Sarah Mayell, Jayesh Mahendra Bhatt, Lutz Naehrlich, Friedrich Pahlke, Martin Wetzke, Alexander Moeller, Matthias Kappler, the chILD-EU collaborators, Nicolaus Schwerk, Elias Seidl

## Abstract

**Background:** Pulmonary fibrosis is of critical importance in childhood interstitial lung disease (chILD), yet fibrosis prevalence, impact on the clinical progression, and survival have not been systematically evaluated.

**Methods:** Data were extracted from the chILD-EU register, a European prospective multicenter cohort study with centralized peer-review on patient inclusion and systematic scoring of computed tomography (CT) scans and lung biopsies. Pulmonary fibrosis was determined based on predefined criteria (fibrosis register) or criteria used in clinical trials (fibrosis trial). We calculated fibrosis rates of chILD entities, evaluated fibrosis criteria and assessed longitudinal pulmonary function testing and survival rates of children with or without fibrosis.

**Results:** 1,071 children diagnosed with chILD were included in the final analysis. The childhood prevalence of fibrosis was for 20.5% (220/1071) according to a single time point, register definition and 11.6% (62/534) according to the dual time point, trial definition. At the age when the children were able to perform pulmonary function tests, those with fibrosis had 15-20% worse predicted forced vital capacity (ppFVC), were older and diagnosed later. Throughout childhood, the disease trajectories assessed as decline in ppFVC and survival did not differ between children with or without pulmonary fibrosis or between the two fibrosis definitions. Overall, survival until the age of 20 years was about 70%.

**Conclusions:** This study assesses the prevalence, pulmonary function progression and survival of pulmonary fibrosis in chILD. The application of standardized criteria for pulmonary fibrosis enables identification of affected children among patients and may support early selection for anti-fibrotic therapies.

## Research in context

### Evidence before this study

Pulmonary fibrosis in childhood interstitial lung disease (chILD) may be responsible for considerable morbidity and mortality, yet its disease-specific prevalence and associated outcome in children remain poorly defined. We searched for studies of children with pulmonary fibrosis published in any language before October 6, 2025, using the search terms: “pulmonary interstitial fibrosis”, “pulmonary fibrosis”, “fibrotic lung disease”, “fibrosing alveolitis” or “fibrosing interstitial lung disease”. Excluding case reports or small series (< 15 children), all solely based on histopathological fibrosis, we identified 2 relevant studies that defined childhood pulmonary fibrosis based on histologic and CT radiologic criteria: One retrospective, single center study on 124 children with diffuse lung disease performed a CT-based analysis and described 19 children with fibrotic features. The other study was a randomized controlled trial enrolling 39 patients into a study of nintedanib. These investigations defined criteria to identify children with pulmonary fibrosis. However, none of these studies used a well-defined, prospectively followed cohort, systematically determined fibrosis in all available lung biopsies or CT scans, assessed long-term pulmonary function testing and survival.

### Added value of this study

To our knowledge, this study is the first comprehensive and prospective analysis of pulmonary fibrosis in chILD. Drawing from the European prospective chILD-EU register, this study includes 1071 children having CT scans and lung biopsies systematically scored. 20.5% had pulmonary fibrosis, based on a single timepoint-register definition, i.e. fibrotic biopsy or one CT scan with fibrosis. Using a dual time point - trial definition, i.e. fibrotic biopsy and one CT or two CT scans in the absence of a biopsy, 11.6% had fibrosis. Children with fibrosis had a 15-20% worse ppFVC at any time point during childhood. In our cohort, overall, survival until the age of 20 years was about 70% and the long-term disease trajectories between children with or without pulmonary fibrosis did not differ throughout childhood.

### Implications of all the available evidence

Lack of standardized criteria for identifying and evaluating pulmonary fibrosis in chILD has led to uncertainty in assessing risk, outcome and treatment planning – especially in the light of emerging anti-fibrotic therapies also for children. This study defined imaging and histological criteria to recognize fibrotic patterns, enabling early identification of patients who may benefit from targeted interventions. The findings provide crucial estimates to inform the design of future clinical trials. Moving forwards, defining and evaluating fibrosis and its progression by clinical teams caring for chILD remains a key objective.

## Background

Childhood interstitial lung disease (chILD) encompasses a diverse group of rare, diffuse parenchymal lung diseases that affect infants, children, and adolescents ^1^. However, little is known about the long-term outcomes of many of these rare conditions. Mortality rates vary by age group with prospective studies reporting rates as high as 13% ^2^. Pulmonary fibrosis in adults as a major cause of death in patients with ILD, has been a major research focus ^3^, leading to international consensus statements from respiratory societies ^4^ and to the development of antifibrotic treatments ^5^. In chILD however, details about pulmonary fibrosis are scarce. A retrospective, single center CT-based study of 124 children with diffuse lung disease identified 19 children with fibrotic features ^6^, and an international review has recently underscored its clinical relevance ^7^. However, no study has yet systematically assessed the prevalence of pulmonary fibrosis in chILD, its impact on disease trajectories or mortality.

A major barrier to advancing this field has been the lack of a standardized definition of pulmonary fibrosis in children. Currently two different definitions are commonly applied, a register-based definition ^7^ and one proposed in the recent first pediatric randomized trial of the antifibrotic agent nintedanib in children ^8^. The significance of these definitions has not been assessed systematically. Both adopted the radiological criteria from adultś definition, including reticulation/linear markings, traction bronchiectasis, architectural distortion and honeycombing ^9^. In children, cystic lucencies are considered as they a hallmark of fibrosing lung disease in early life, particularly in genetically defined surfactant dysfunction disorders ^10^. Although longitudinal data in children are scarce, cystic lesions have been shown to increase in number and size over time in disorders such as surfactant protein C deficiency and adenosine triphosphate-binding cassette subfamily A member 3 (ABCA3) deficiency ^11–14^. However, the impact of including cystic (parenchymal) lesions as a defining criterion for fibrosis has not been formally studied.

Notably, fibrosing processes in chILD affect lungs that are still undergoing rapid development and have a substantial regenerative potential, extending well into adolescence ^15^. This plasticity offers a unique potential for reversibility of even fibrotic processes, similar to children with bronchiectasis ^16^. These developmental differences emphasize the need for pediatric specific criteria and interpretation when evaluating fibrosis in chILD.

The aim of this study was to determine the prevalence of pulmonary fibrosis and its impact on lung function as well as survival in a large, prospectively followed cohort of children, using both the trial and registry fibrosis definitions. Additionally, we aimed to explore the role and clinical significance of cystic (parenchymal) changes in paediatric fibrosing interstitial lung disease.

## Methods

### Study design, participants and procedures

This study was based on prospectively collected data from the chILD-EU register, a multi-center European study (www.childeu.net). Participating centers adhered to all contractual, legal, and ethical standards before enrolling patients. Each patient and/or caregiver provided age-appropriate verbal assent and written informed consent. The study was approved by the lead Ethics committee of the University Hospital Munich (20-0329, 22-0133) and all local committees. We followed the Declaration of Helsinki and STROBE reporting guidelines to ensure completeness and transparency.

Children suspected of having chILD were identified by their treating physicians and submitted to a web-based platform for peer review. Diagnostic categorization was based on systematically obtained data ^17^. A multidisciplinary team including clinicians, radiologists, and pathologists with expertise in chILD established the diagnosis in accordance with the guidelines of the American Thoracic Society ^18^ and the European management platform for childhood interstitial lung diseases ^19^. Diagnoses were subsequently grouped into predefined categories ^20,21^. Patients with airway disorders, pleural disorders, non-ILD infections, gross structural abnormalities or neoplasms were excluded. Participants were prospectively followed up at 6 and 12 months and then annually. To minimize loss to follow-up, the registry staff contacted study sites via the online platform, emails, and telephone at least four times per year. Detailed information on the implementation and operation of the chILD-EU register is published elsewhere ^17^. CT imaging was systematically reviewed by three experienced radiologists JL-Z (thoracic radiologist, 10-years pediatric-board certified), BK (23-years pediatric-board certified), IK-S (12-years pediatric board certified)) using a consensus-based approach. Each scan was evaluated for the presence or absence of reticulation/linear markings, traction bronchiectasis, architectural distortion, honeycombing or cystic (parenchymal) lesions, as defined by the Fleischner Society ^22^.

Lung biopsies were assessed by a pathologist (SR, (14-years board certified, 12-years specialized in pediatric lung pathology)) evaluating for interstitial fibrosis, fibroblastic foci, or honeycombing. These scorings were extracted and secondary data analysis was performed based on pre-set criteria to determine if fibrosis is present (Fig. 1).

**Fig. 1.**
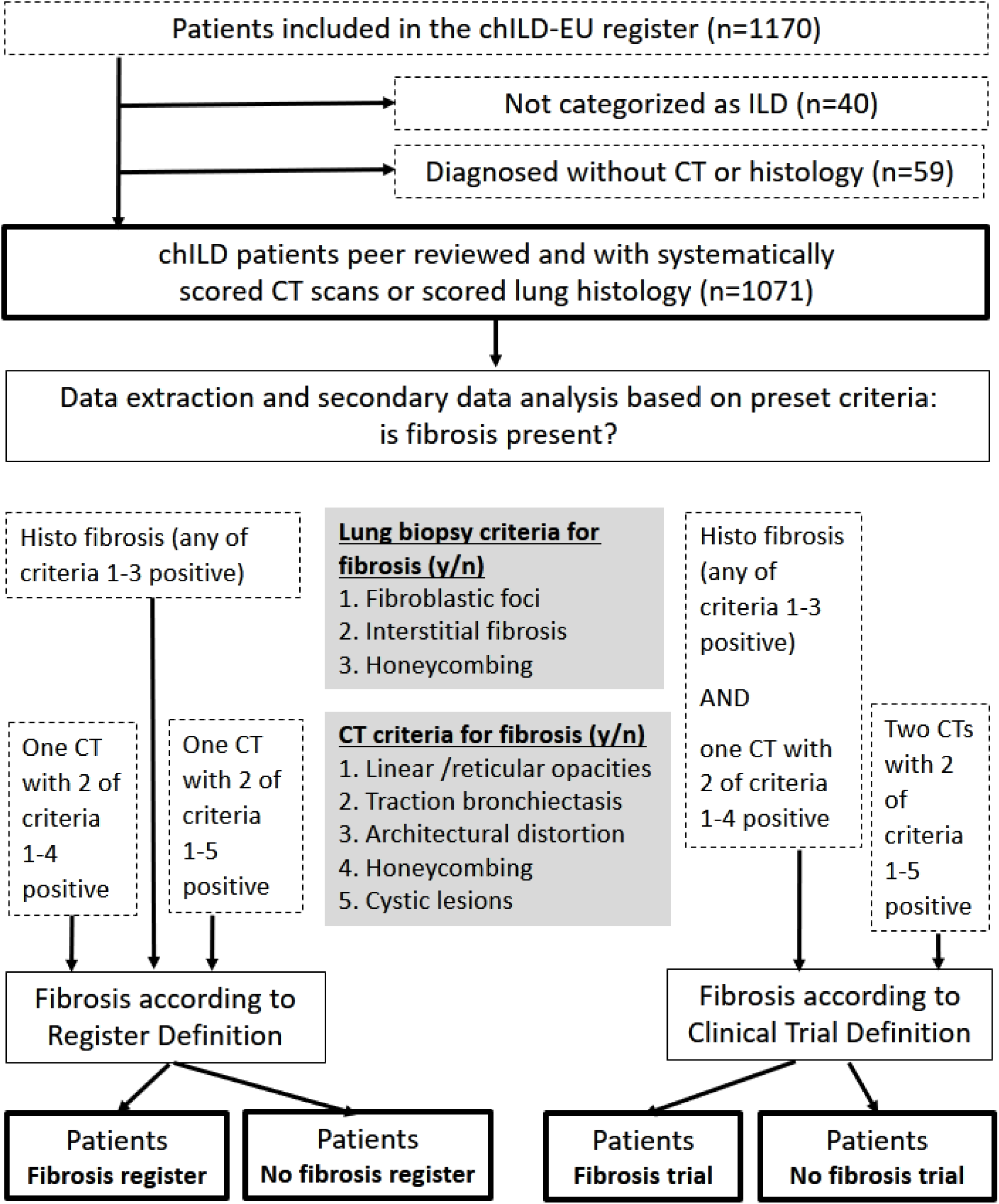
Diagram (CONSORT) of study design, definitions of fibrosis and participants included.

### Outcomes

The primary outcome was the presence or absence of fibrosing lung disease. The fibrosis definition used in the register (“Fibrosis register”)(Fig. 1, left side) included patients if at least CT scan showed 2 of the four criteria “linear/reticular opacities”, “traction bronchiectasis”, “architectural distortion”, and “honeycombing” positive or showed 2 of the five criteria i.e. the previous four criteria plus “cystic lesions” positive or if the lung biopsy showed “interstitial fibrosis”, “fibroblastic foci” or “honeycombing”.

The fibrosis definition used in the trial (“Fibrosis trial”)(Fig. 1, right side) included patients if at least two CT scans showed 2 of the five criteria “linear/reticular opacities”, “traction bronchiectasis”, “architectural distortion”, “honeycombing” and “cystic lesions” positive or if at least one CT scan had two of the four criteria “linear or reticular opacities”, “traction bronchiectasis”, “architectural distortion”, and “honeycombing” positive and had a lung biopsy with “interstitial fibrosis”, “fibroblastic foci” or “honeycombing”.

Clinical and socio-demographic variables were retrieved from chILD EU register (Tables 1 and 2), including pulmonary function parameters forced expiratory volume in one second (FEV1), forced vital capacity (FVC) and maximum mid expiratory flow (MMEF) expressed as percent predicted (pp) using the GLI equations ^17^, Fan severity score ^23^, BMI z-scores (www.cdc.gov/growth-chart-training), failure to thrive and respiratory rate z-score ^24^. Survival was defined as the time from birth to age at death or lung transplantation. All clinical variables were entered into an electronic case reporting system completed by the responsible site investigators.

**Table 1.**
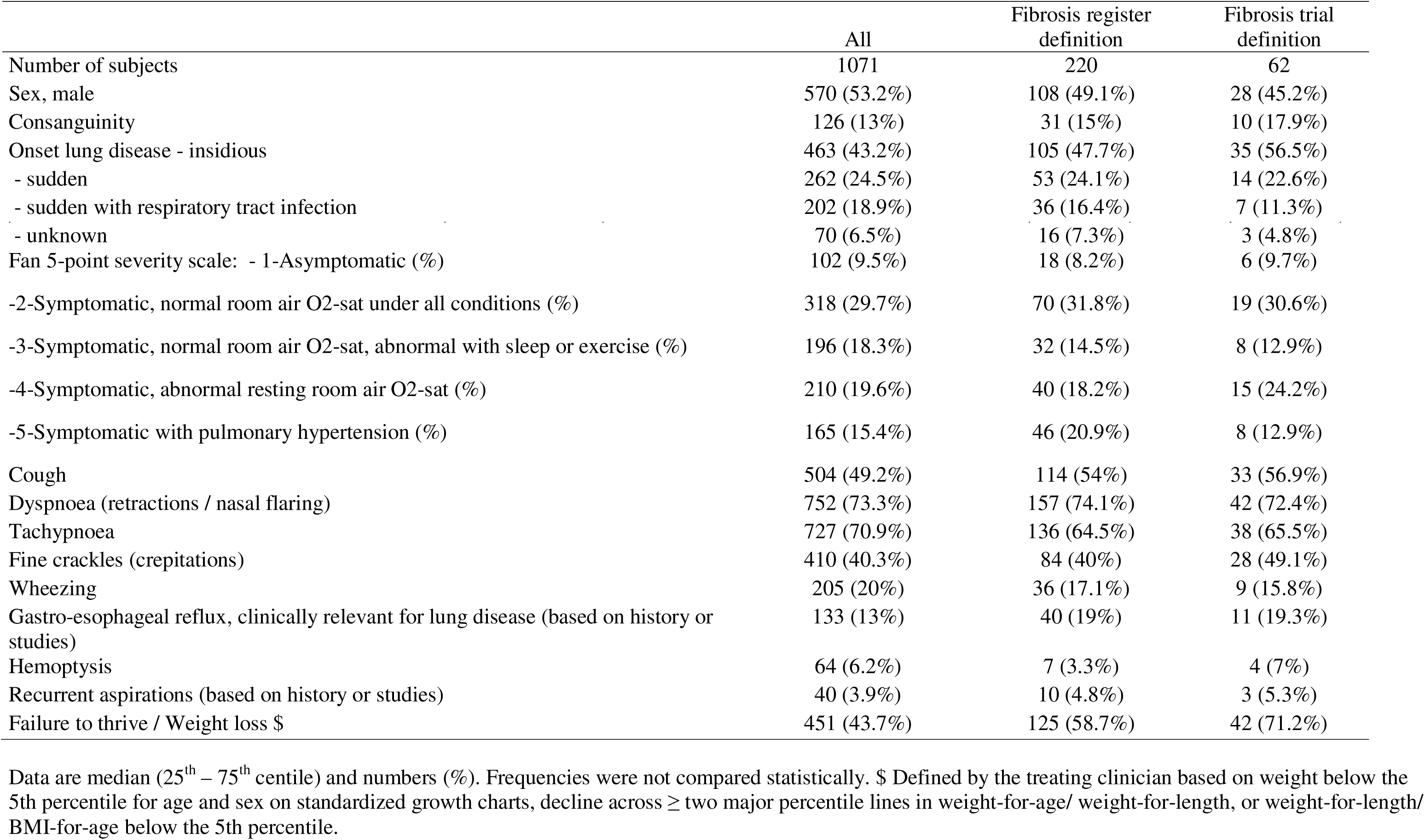
Demographics and medical history at inclusion into the register.

|  | All | Fibrosis register<br>definition | Fibrosis trial<br>definition |
| --- | --- | --- | --- |
| Number of subjects | 1071 | 220 | 62 |
| Sex, male | 570 (53.2%) | 108 (49.1%) | 28 (45.2%) |
| Consanguinity | 126 (13%) | 31 (15%) | 10 (17.9%) |
| Onset lung disease - insidious | 463 (43.2%) | 105 (47.7%) | 35 (56.5%) |
| - sudden | 262 (24.5%) | 53 (24.1%) | 14 (22.6%) |
| - sudden with respiratory tract infection | 202 (18.9%) | 36 (16.4%) | 7 (11.3%) |
| - unknown | 70 (6.5%) | 16 (7.3%) | 3 (4.8%) |
| Fan 5-point severity scale: - 1-Asymptomatic (%) | 102 (9.5%) | 18 (8.2%) | 6 (9.7%) |
| -2-Symptomatic, normal room air O2-sat under all conditions (%) | 318 (29.7%) | 70 (31.8%) | 19 (30.6%) |
| -3-Symptomatic, normal room air O2-sat, abnormal with sleep or exercise (%) | 196 (18.3%) | 32 (14.5%) | 8 (12.9%) |
| -4-Symptomatic, abnormal resting room air O2-sat (%) | 210 (19.6%) | 40 (18.2%) | 15 (24.2%) |
| -5-Symptomatic with pulmonary hypertension (%) | 165 (15.4%) | 46 (20.9%) | 8 (12.9%) |
| Cough | 504 (49.2%) | 114 (54%) | 33 (56.9%) |
| Dyspnoea (retractions / nasal flaring) | 752 (73.3%) | 157 (74.1%) | 42 (72.4%) |
| Tachypnoea | 727 (70.9%) | 136 (64.5%) | 38 (65.5%) |
| Fine crackles (crepitations) | 410 (40.3%) | 84 (40%) | 28 (49.1%) |
| Wheezing | 205 (20%) | 36 (17.1%) | 9 (15.8%) |
| Gastro-esophageal reflux, clinically relevant for lung disease (based on history or studies) | 133 (13%) | 40 (19%) | 11 (19.3%) |
| Hemoptysis | 64 (6.2%) | 7 (3.3%) | 4 (7%) |
| Recurrent aspirations (based on history or studies) | 40 (3.9%) | 10 (4.8%) | 3 (5.3%) |
| Failure to thrive / Weight loss \$ | 451 (43.7%) | 125 (58.7%) | 42 (71.2%) |

Data are median (25<sup>th</sup> – 75<sup>th</sup> centile) and numbers (%). Frequencies were not compared statistically. \$ Defined by the treating clinician based on weight below the 5th percentile for age and sex on standardized growth charts, decline across ≥ two major percentile lines in weight-for-age/ weight-for-length, or weight-for-length/ BMI-for-age below the 5th percentile.

### Statistics

Data analysis was performed using SPSS statistical analysis software (Version 26.0), R Project for Statistical Computing (Version 4.4.2) and Graph Pad Prism (Version 8.3.0). Demographic data and clinical features were reported as numbers (percentage) or median (lower and upper quartile) and number of subjects included. Group comparisons were conducted using Mann-Whitney test and correction of multiple comparisons. Longitudinal changes in ppFVC were assessed using linear mixed-effects models. Fixed effects included fibrosis status (Yes/No), yearly visit (categorical), baseline PFT (0Y) age (age when first pulmonary function test (PFT) was available), and the interaction between fibrosis status and visit. A random intercept for each patient was specified to account for within-subject variability. Residual correlation was modelled using a first-order autoregressive (AR1) structure, and heteroscedasticity across visits was modelled with a variance structure. Missing data were assumed to be missing at random. Estimated marginal means were obtained for each group at each visit and were used to compare fibrosis groups at each visit and to examine within-group changes across visits, including changes from baseline PFT (0Y). Pairwise comparisons of estimated marginal means were adjusted for multiple testing. For within-group comparisons across visits, Tukey’s method was applied. For comparisons between fibrosis groups at each visit, p-values were adjusted across visits using the Holm procedure. Statistical significance was defined as two-sided p < 0.05.

### Role of the funding source

The chILD-EU register is a collaborative study (Trial registration number: NCT02852928) originally funded by the FP7 project 305653-chILD-EU (www.childeu.net) and sustained by participating institutions. Relevant funding for this study was provided by the Deutsche Forschungsgemeinschaft (DFG, Gr970/9-1 and 9-2) and Boehringer-Ingelheim, Germany. The funders had no role in study design, conduct or interpretation, nor in writing or submitting the manuscript for publication.

## Results

### Study population

1071 children from 23 European countries were included in the final analysis (Figure 1, Supplemental Table 1), representing the entire spectrum of chILD conditions. At inclusion into the register, this large cohort had the characteristic features of subjects with chILD, including a small preponderance of males, 13% consanguinity, a mainly insidious disease onset (43.2%), a broad distribution of disease severity, typical symptoms such as dyspnea (73.3%), fine crackles (40.3%) and failure to thrive (43.7%) (Table 1). The median age at inclusion was 2.4 years, chronic lung disease symptoms started at 0.4 years of age and ILD was diagnosed at 0.8 years of age. Lung function testing data, in those children old enough to perform, respiratory rate and BMI were substantially decreased and listed in Table 2.

**Table 2.** Age at various time points and quantitative respiratory and general data at inclusion into the register.

|  | All | Fibrosis according to register definition |  | Fibrosis according to trial definition |  |
| --- | --- | --- | --- | --- | --- |
|  |  | yes | no | yes | no |
| Age (years) | 2.35 (0.6; 9.18) 1070\$ | 7.2 (1.3; 13.02) 220*** | 1.75 (0.5; 7.9) 850 | 9.25 (4.08; 13.75) 62### | 4 (0.8; 10.2) 472 |
| Age at start of chronic lung disease symptoms (years) | 0.4 (0; 4.2) 1004 | 1 (0; 8.05) 211* | 0.4 (0; 3.2) 793 | 1 (0; 8.15) 59 | 0.6 (0; 6.3) 451 |
| Age at first ILD detection (years) | 0.8 (0.3; 6.25) 1011 | 2.85 (0.3; 9.9) 212*** | 0.7 (0.25; 4.7) 799 | 4.4 (0.25; 10.05) 59 | 1.2 (0.3; 7.8) 452 |
| Age at biopsy (years) | 2.1 (0.5; 9) 485 | 4.3 (0.8; 10.62) 152*** | 1.5 (0.4; 7.5) 333 | 5.75 (1.12; 10.22) 42 | 2.05 (0.5; 9.2) 314 |
| Age at LTX (years) | 10 (3.9; 13.82) 52 | 7.65 (3.12; 12.28) 20 | 10 (4.3; 14.48) 32 | 10.5 (10.3; 11.9) 5 | 10.15 (4.18; 13.92) 32 |
| Age at death (years) | 1.2 (0.2; 7.25) 123 | 8.55 (1.08; 16.22) 36*** | 0.6 (0.1; 4.05) 87 | 12.9 (8.55; 18.2) 15### | 2.8 (0.45; 6.6) 54 |
| ppFEV1 | 65 (44; 79.5) 304 | 51 (39; 70) 89*** | 68 (49.5; 85) 215 | 48 (33; 65) 29## | 67 (43.25; 84) 170 |
| ppFVC | 67.5 (47; 83) 306 | 55 (39; 72.5) 91*** | 73 (54.5; 87) 215 | 51.5 (43.25; 65) 30### | 72.5 (49; 87) 170 |
| ppMEF25/75 | 58 (35; 90) 241 | 49 (28; 74) 69 | 65 (42; 93.25) 172 | 48.5 (25.75; 72.75) 26 | 63.5 (40; 92) 130 |
| ppDLCO | 61 (42; 80) 89 | 42 (38; 61) 29*** | 68.5 (54; 89.25) 60 | 49.5 (39.75; 59.5) 8 | 68.5 (47.25; 93.75) 50 |
| Respiratory rate z-score | 2.56 (0.44; 4.18) 500 | 3.34 (0.9; 5.09) 104* | 2.44 (0.42; 4.18) 396 | 3.88 (1.59; 7.45) 28 | 2.66 (0.52; 4.95) 206 |
| BMI z-score | -0.9 (-2.13; 0.28) 970 | -1.37 (-2.37; -0.03) 205* | -0.81 (-2.06; 0.32) 765 | -1.31 (-2.66; -0.1) 55 | -0.81 (-2.08; 0.36) 439 |

Data are median (25<sup>th</sup> – 75<sup>th</sup> quartile) and numbers of subjects. \$ exact age at inclusion into the register in one case unknown. Comparisons were made by Mann-Whitney test and P was corrected for the 2 comparisons indicated. Fibrosis register \*\*\* P<0.002, \* P<0.05, Fibrosis trial; ### P<0.002, ## P<0.004. pp = % predicted.

Of the 1,071 chILD patients who had at least one CT or lung histology, 220 (20.5%) had fibrosis according to the register definition criteria. Among the 534 subjects who had at least two CTs or one CT and a lung histology, 62 (11.6%) children had pulmonary fibrosis according to the trial definition of fibrosis (Table 2). At inclusion into the register, patients from both fibrosis groups when compared to the respective patients with no fibrosis, were older, had worse initial ppFVC and ppFEV1, and had a higher age at death. Compared to overall average, fibrosis patients tended to be more often female, had a higher rate of consanguinity, more frequently an insidious start of their disease, more coughing, more gastro-esophageal reflux and recurrent aspirations and much more frequently failure to thrive or weight loss; BMI z-score was lower in patients with register defined fibrosis (Table 1). Compared with the overall cohort, patients with pulmonary fibrosis were more often female and had a higher rate of consanguinity. They more frequently presented with an insidious disease onset, cough, gastro-oesophageal reflux, recurrent aspirations, and, notably, failure to thrive or weight loss. The BMI z-score was lower in patients with registry-defined fibrosis (Table 1).

### The criteria to diagnose fibrosis in chILD

Among the group of 220 patients with fibrosis according to register definition, linear or reticulation opacities were present in 73.2%, followed by cystic lesions (51.4%), architectural distortion (38.6%), traction bronchiectasis (28.6%), and honey combing (9.5%)(Table 3). Among patients who did not meet the criteria for pulmonary fibrosis, linear/ reticular opacities were present in 16.8%, cystic lesions in 7.8%, and all other features in less than 2% (Table 3).

**Table 3.**
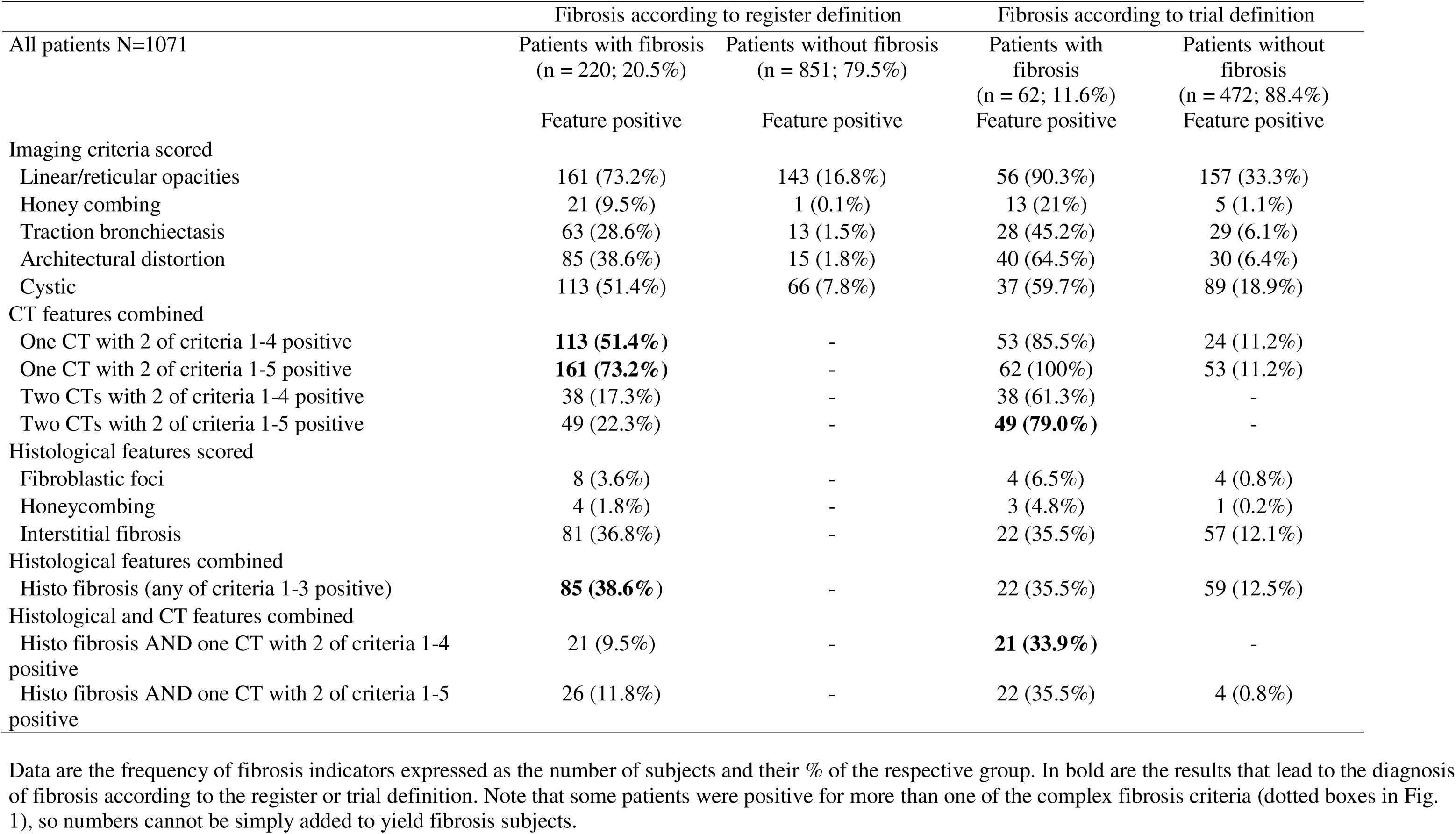
Criteria fulfilled to diagnose fibrosing lung disease and the role of cystic lesions on CT scans.

| All patients N=1071 | Fibrosis according to register definition |  | Fibrosis according to trial definition |  |
| --- | --- | --- | --- | --- |
|  | Patients with fibrosis<br>(n = 220; 20.5%) | Patients without fibrosis<br>(n = 851; 79.5%) | Patients with<br>fibrosis<br>(n = 62; 11.6%) | Patients without<br>fibrosis<br>(n = 472; 88.4%) |
|  | Feature positive | Feature positive | Feature positive | Feature positive |
| Imaging criteria scored |  |  |  |  |
| Linear/reticular opacities | 161 (73.2%) | 143 (16.8%) | 56 (90.3%) | 157 (33.3%) |
| Honey combing | 21 (9.5%) | 1 (0.1%) | 13 (21%) | 5 (1.1%) |
| Traction bronchiectasis | 63 (28.6%) | 13 (1.5%) | 28 (45.2%) | 29 (6.1%) |
| Architectural distortion | 85 (38.6%) | 15 (1.8%) | 40 (64.5%) | 30 (6.4%) |
| Cystic | 113 (51.4%) | 66 (7.8%) | 37 (59.7%) | 89 (18.9%) |
| CT features combined |  |  |  |  |
| One CT with 2 of criteria 1-4 positive | **113 (51.4%)** | - | 53 (85.5%) | 24 (11.2%) |
| One CT with 2 of criteria 1-5 positive | **161 (73.2%)** | - | 62 (100%) | 53 (11.2%) |
| Two CTs with 2 of criteria 1-4 positive | 38 (17.3%) | - | 38 (61.3%) | - |
| Two CTs with 2 of criteria 1-5 positive | 49 (22.3%) | - | **49 (79.0%)** | - |
| Histological features scored |  |  |  |  |
| Fibroblastic foci | 8 (3.6%) | - | 4 (6.5%) | 4 (0.8%) |
| Honeycombing | 4 (1.8%) | - | 3 (4.8%) | 1 (0.2%) |
| Interstitial fibrosis | 81 (36.8%) | - | 22 (35.5%) | 57 (12.1%) |
| Histological features combined |  |  |  |  |
| Histo fibrosis (any of criteria 1-3 positive) | **85 (38.6%)** | - | 22 (35.5%) | 59 (12.5%) |
| Histological and CT features combined |  |  |  |  |
| Histo fibrosis AND one CT with 2 of criteria 1-4 positive | 21 (9.5%) | - | **21 (33.9%)** | - |
| Histo fibrosis AND one CT with 2 of criteria 1-5 positive | 26 (11.8%) | - | 22 (35.5%) | 4 (0.8%) |

Of the subjects meeting the diagnostic criteria for pulmonary fibrosis, 73.2% were diagnosed based on CT findings alone, i.e. 51.4% based on the criteria linear/reticulation, honey combing, traction bronchiectasis and architectural distortion; additional 21.8% were diagnosed considering also cystic CT lesions. Histology alone added the missing 26.8% fibrosis cases (Table 3).

Among the 534 patients with 2 CT scans or one CT and a histology available, 62 patients fulfilled the trial fibrosis definition. Of these subjects, 79% were diagnosed based on CT findings. Histological evaluation of lung tissue in combination with a CT scan accounted for the remaining 21% of fibrosis diagnoses (Table 3). The frequency of single radiological fibrotic features in patients not meeting the necessary criteria for pulmonary fibrosis was high, ranging between 33.3% (linear/reticulation) and 1.1% (honey combing)(Table 3). The strict trial definition of fibrosis resulted in 5.8% of the patients defined as fibrotic, compared to 20.5%, when the register definition was used.

Considering CTs alone and the usage of the “cystic criterion” increased fibrosis diagnosis from 61.3 to 79.0% of included subjects (+17.7%), using the trial criteria, and from 51.4 to 73.2% (+21.8%) using the register definition of pulmonary fibrosis. To assess the impact of cystic lesions on outcome, we compared lung function testing results in patients with and without cystic (parenchymal) lesions. Among all patients, those with cysts (median ppFVC 65.5% (IQR 48.5%-79.0%), n = 108) had a worse ppFVC than those without (70.1% (52.0%-91.0%), n = 430; p = 0.0253, Wilcoxon test). Among patients with fibrosis according to register definition and having just two fibrosis criteria on their CT scans, subjects with cystic (parenchymal) lesions (n=31) had higher baseline PFT (0Y) ppFVC values (median ppFVC 70.0% (IQR 63.0-93.5) compared to those without (52.0% (24.5-56.5), n=27, P < 0.0001, Wilcoxon test). The initial difference in ppFVC between the groups was no longer evident after one year and remained absent throughout the 8-year follow-up (Supplemental Figure 1).

### The frequency of fibrosing lung disease at the diagnosis and subcategory level

Fibrosis rates for each diagnosis were determined and classified into three categories: conditions affecting the lungs only, conditions associated with systemic diseases, and those due to external exposures (Table 4). Among the first, ABCA3 deficiency, SFTPC dysfunction disorders, nonspecific interstitial pneumonia, lymphocytic interstitial pneumonia and capillary hemangiomatosis had frequencies of fibrosing lung disease in more than 20%. Importantly, conditions associated with systemic diseases often had fibrosis frequencies above 20%, and up to above 50% in aminoacyl-tRNA synthetase (ARS)-deficiency, COPA syndrome, TBX4-associated disease and infantile-onset STING-associated vasculopathy. Lastly, conditions related to exogenous injury, in particular chronic lung disease of prematurity (BPD-cLD), also presented with lung fibrosis (Table 4).

**Table 4.**
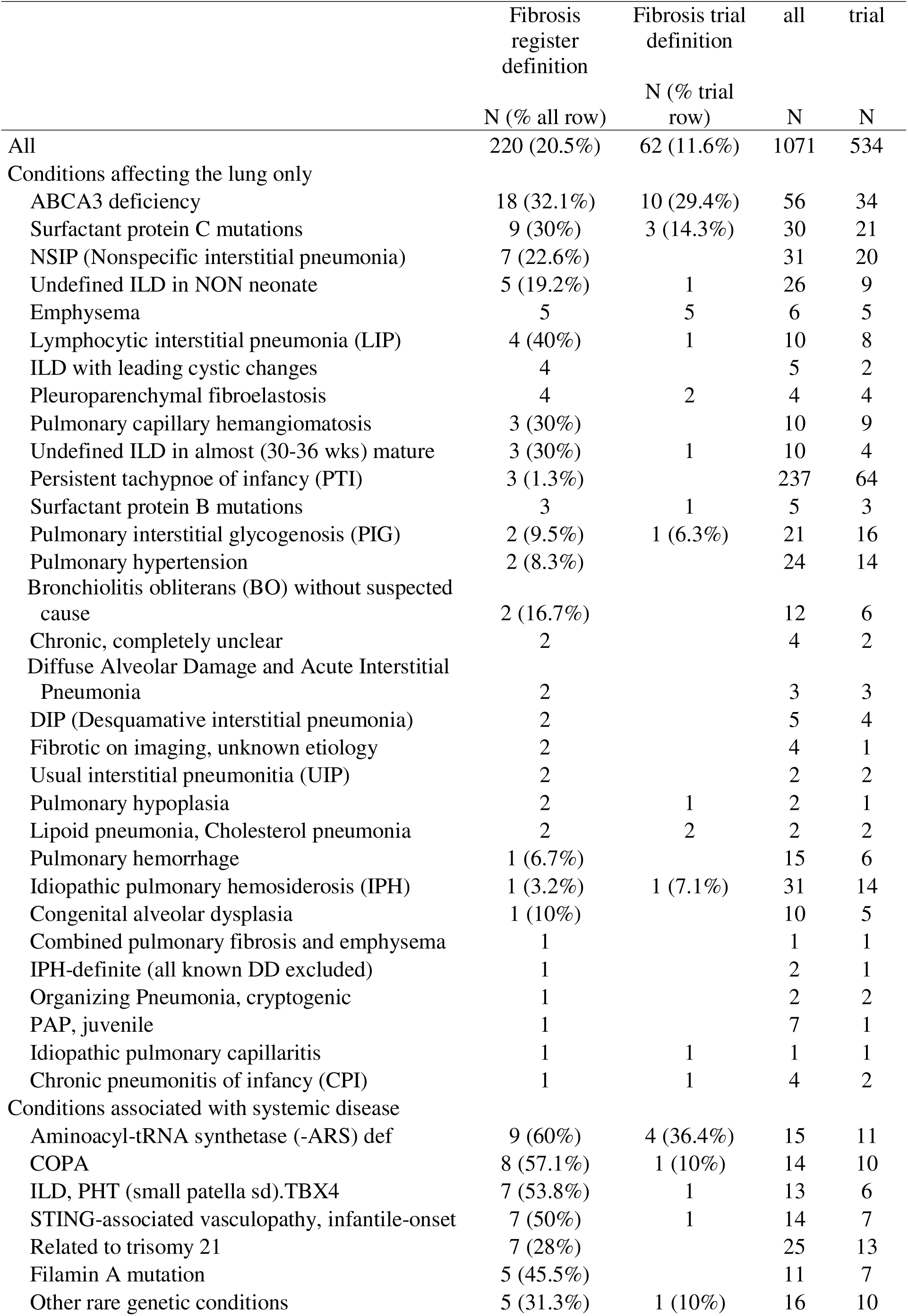
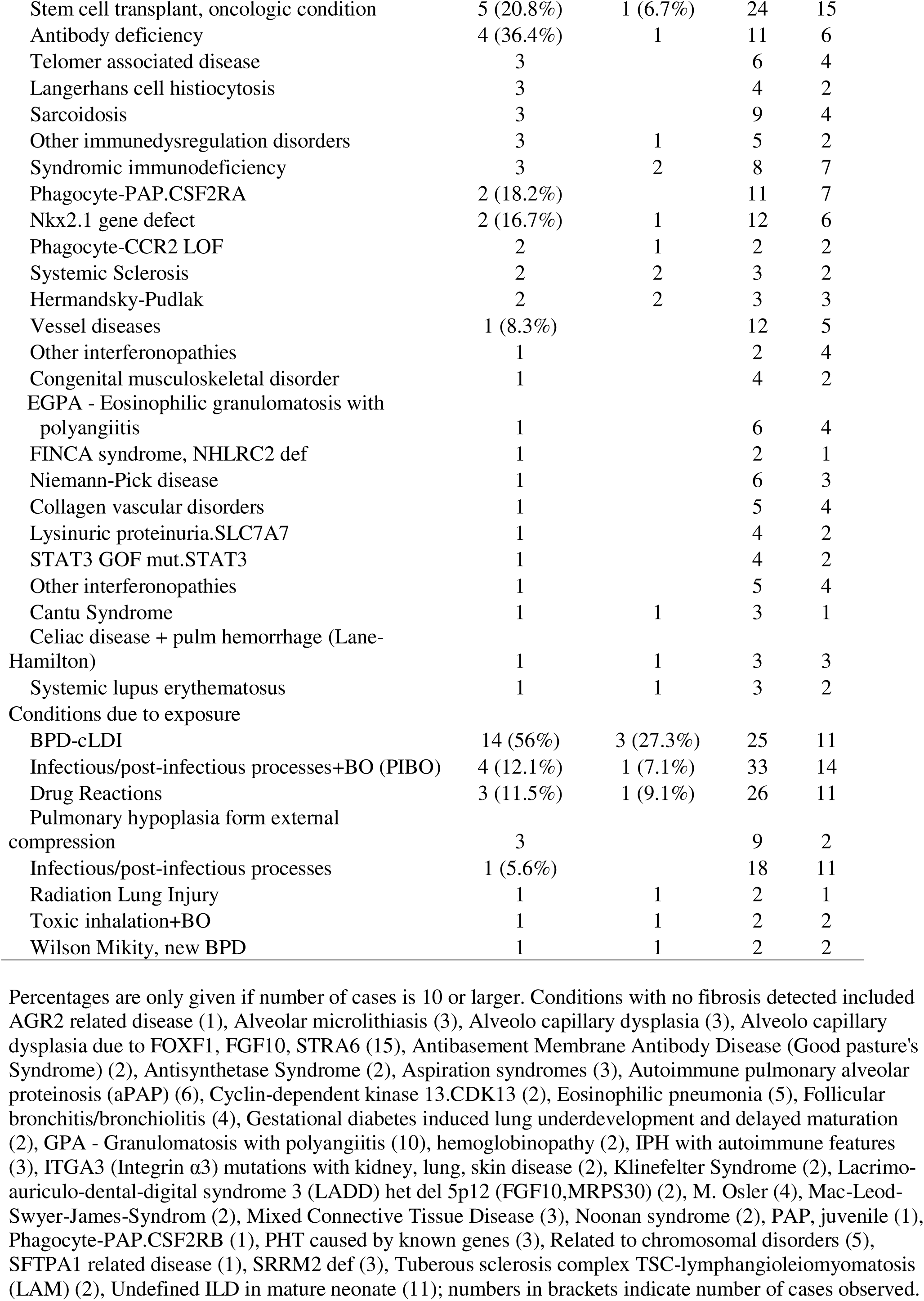
Frequency of pulmonary fibrosis according to disease category and fibrosis definition.

|  | Fibrosis<br>register<br>definition | Fibrosis trial<br>definition | all | trial |
| --- | --- | --- | --- | --- |
|  | N (% all row) | N (% trial<br>row) | N | N |
| All | 220 (20.5%) | 62 (11.6%) | 1071 | 534 |
| Conditions affecting the lung only |  |  |  |  |
| ABCA3 deficiency | 18 (32.1%) | 10 (29.4%) | 56 | 34 |
| Surfactant protein C mutations | 9 (30%) | 3 (14.3%) | 30 | 21 |
| NSIP (Nonspecific interstitial pneumonia) | 7 (22.6%) |  | 31 | 20 |
| Undefined ILD in NON neonate | 5 (19.2%) | 1 | 26 | 9 |
| Emphysema | 5 | 5 | 6 | 5 |
| Lymphocytic interstitial pneumonia (LIP) | 4 (40%) | 1 | 10 | 8 |
| ILD with leading cystic changes | 4 |  | 5 | 2 |
| Pleuroparenchymal fibroelastosis | 4 | 2 | 4 | 4 |
| Pulmonary capillary hemangiomatosis | 3 (30%) |  | 10 | 9 |
| Undefined ILD in almost (30-36 wks) mature | 3 (30%) | 1 | 10 | 4 |
| Persistent tachypnoe of infancy (PTI) | 3 (1.3%) |  | 237 | 64 |
| Surfactant protein B mutations | 3 | 1 | 5 | 3 |
| Pulmonary interstitial glycogenosis (PIG) | 2 (9.5%) | 1 (6.3%) | 21 | 16 |
| Pulmonary hypertension | 2 (8.3%) |  | 24 | 14 |
| Bronchiolitis obliterans (BO) without suspected<br>cause | 2 (16.7%) |  | 12 | 6 |
| Chronic, completely unclear | 2 |  | 4 | 2 |
| Diffuse Alveolar Damage and Acute Interstitial<br>Pneumonia | 2 |  | 3 | 3 |
| DIP (Desquamative interstitial pneumonia) | 2 |  | 5 | 4 |
| Fibrotic on imaging, unknown etiology | 2 |  | 4 | 1 |
| Usual interstitial pneumonitis (UIP) | 2 |  | 2 | 2 |
| Pulmonary hypoplasia | 2 | 1 | 2 | 1 |
| Lipoid pneumonia, Cholesterol pneumonia | 2 | 2 | 2 | 2 |
| Pulmonary hemorrhage | 1 (6.7%) |  | 15 | 6 |
| Idiopathic pulmonary hemosiderosis (IPH) | 1 (3.2%) | 1 (7.1%) | 31 | 14 |
| Congenital alveolar dysplasia | 1 (10%) |  | 10 | 5 |
| Combined pulmonary fibrosis and emphysema | 1 |  | 1 | 1 |
| IPH-definite (all known DD excluded) | 1 |  | 2 | 1 |
| Organizing Pneumonia, cryptogenic | 1 |  | 2 | 2 |
| PAP, juvenile | 1 |  | 7 | 1 |
| Idiopathic pulmonary capillaritis | 1 | 1 | 1 | 1 |
| Chronic pneumonitis of infancy (CPI) | 1 | 1 | 4 | 2 |
| Conditions associated with systemic disease |  |  |  |  |
| Aminoacyl-tRNA synthetase (-ARS) def | 9 (60%) | 4 (36.4%) | 15 | 11 |
| COPA | 8 (57.1%) | 1 (10%) | 14 | 10 |
| ILD, PHT (small patella sd).TBX4 | 7 (53.8%) | 1 | 13 | 6 |
| STING-associated vasculopathy, infantile-onset | 7 (50%) | 1 | 14 | 7 |
| Related to trisomy 21 | 7 (28%) |  | 25 | 13 |
| Filamin A mutation | 5 (45.5%) |  | 11 | 7 |
| Other rare genetic conditions | 5 (31.3%) | 1 (10%) | 16 | 10 |
| Stem cell transplant, oncologic condition | 5 (20.8%) | 1 (6.7%) | 24 | 15 |
| Antibody deficiency | 4 (36.4%) | 1 | 11 | 6 |
| Telomer associated disease | 3 |  | 6 | 4 |
| Langerhans cell histiocytosis | 3 |  | 4 | 2 |
| Sarcoidosis | 3 |  | 9 | 4 |
| Other immunedysregulation disorders | 3 | 1 | 5 | 2 |
| Syndromic immunodeficiency | 3 | 2 | 8 | 7 |
| Phagocyte-PAP.CSF2RA | 2 (18.2%) |  | 11 | 7 |
| Nkx2.1 gene defect | 2 (16.7%) | 1 | 12 | 6 |
| Phagocyte-CCR2 LOF | 2 | 1 | 2 | 2 |
| Systemic Sclerosis | 2 | 2 | 3 | 2 |
| Hermandsky-Pudlak | 2 | 2 | 3 | 3 |
| Vessel diseases | 1 (8.3%) |  | 12 | 5 |
| Other interferonopathies | 1 |  | 2 | 4 |
| Congenital musculoskeletal disorder | 1 |  | 4 | 2 |
| EGPA - Eosinophilic granulomatosis with polyangiitis | 1 |  | 6 | 4 |
| FINCA syndrome, NHLRC2 def | 1 |  | 2 | 1 |
| Niemann-Pick disease | 1 |  | 6 | 3 |
| Collagen vascular disorders | 1 |  | 5 | 4 |
| Lysinuric proteinuria.SLC7A7 | 1 |  | 4 | 2 |
| STAT3 GOF mut.STAT3 | 1 |  | 4 | 2 |
| Other interferonopathies | 1 |  | 5 | 4 |
| Cantu Syndrome | 1 | 1 | 3 | 1 |
| Celiac disease + pulm hemorrhage (Lane-Hamilton) | 1 | 1 | 3 | 3 |
| Systemic lupus erythematosus | 1 | 1 | 3 | 2 |
| Conditions due to exposure |  |  |  |  |
| BPD-cLDI | 14 (56%) | 3 (27.3%) | 25 | 11 |
| Infectious/post-infectious processes+BO (PIBO) | 4 (12.1%) | 1 (7.1%) | 33 | 14 |
| Drug Reactions | 3 (11.5%) | 1 (9.1%) | 26 | 11 |
| Pulmonary hypoplasia form external compression | 3 |  | 9 | 2 |
| Infectious/post-infectious processes | 1 (5.6%) |  | 18 | 11 |
| Radiation Lung Injury | 1 | 1 | 2 | 1 |
| Toxic inhalation+BO | 1 | 1 | 2 | 2 |
| Wilson Mikity, new BPD | 1 | 1 | 2 | 2 |

Percentages are only given if number of cases is 10 or larger. Conditions with no fibrosis detected included AGR2 related disease (1), Alveolar microlithiasis (3), Alveolo capillary dysplasia (3), Alveolo capillary dysplasia due to FOXF1, FGF10, STRA6 (15), Antibasement Membrane Antibody Disease (Good pasture's Syndrome) (2), Antisynthetase Syndrome (2), Aspiration syndromes (3), Autoimmune pulmonary alveolar proteinosis (aPAP) (6), Cyclin-dependent kinase 13.CDK13 (2), Eosinophilic pneumonia (5), Follicular bronchitis/bronchiolitis (4), Gestational diabetes induced lung underdevelopment and delayed maturation (2), GPA - Granulomatosis with polyangiitis (10), hemoglobinopathy (2), IPH with autoimmune features (3), ITGA3 (Integrin $\alpha 3$ ) mutations with kidney, lung, skin disease (2), Klinefelter Syndrome (2), Lacrimo-auriculo-dental-digital syndrome 3 (LADD) het del 5p12 (FGF10,MRPS30) (2), M. Osler (4), Mac-Leod-Swyer-James-Syndrom (2), Mixed Connective Tissue Disease (3), Noonan syndrome (2), PAP, juvenile (1), Phagocyte-PAP.CSF2RB (1), PHT caused by known genes (3), Related to chromosomal disorders (5), SFTPA1 related disease (1), SRRM2 def (3), Tuberous sclerosis complex TSC-lymphangioliomyomatosis (LAM) (2), Undefined ILD in mature neonate (11); numbers in brackets indicate number of cases observed.

### Longitudinal pulmonary function trajectories

Using the registry definition of fibrosis, the linear mixed-effects model estimated consistently lower ppFVC in children with pulmonary fibrosis compared with those without (Figure 2). At baseline PFT (0Y) (age 7.5 (5.3 to 11.8 years), the estimated marginal mean ppFVC was 73.3% (95% CI: 70.8,75.7) (age 10.3 (6.6 to 14.4 years) in the non-fibrosis group and 59.8% (95% CI: 55.5,64.1) in the fibrosis group, corresponding to a mean difference of −13.4% (95% CI: −18.4, −8.5; p<0.0001). These differences remained statistically significant at all follow-up visits, ranging from −13.3% to −22.0% over 1–8Y (all adjusted p<0.0001; Supplemental Table 2). Within-group trajectories showed improvement in the non-fibrosis group but not in the fibrosis group. In the non-fibrosis group, ppFVC increased from baseline PFT (0Y) by 3.7–8.4% during 1–4Y (all adjusted p≤0.0082) and remained 6.9–7.5% above baseline PFT (0Y) during 5–8Y (all adjusted p≤0.0001; Supplemental Table 3). In contrast, the fibrosis group showed no significant within-group differences between any visits (all adjusted p>0.05; Supplemental Table 3).

**Fig. 2.**
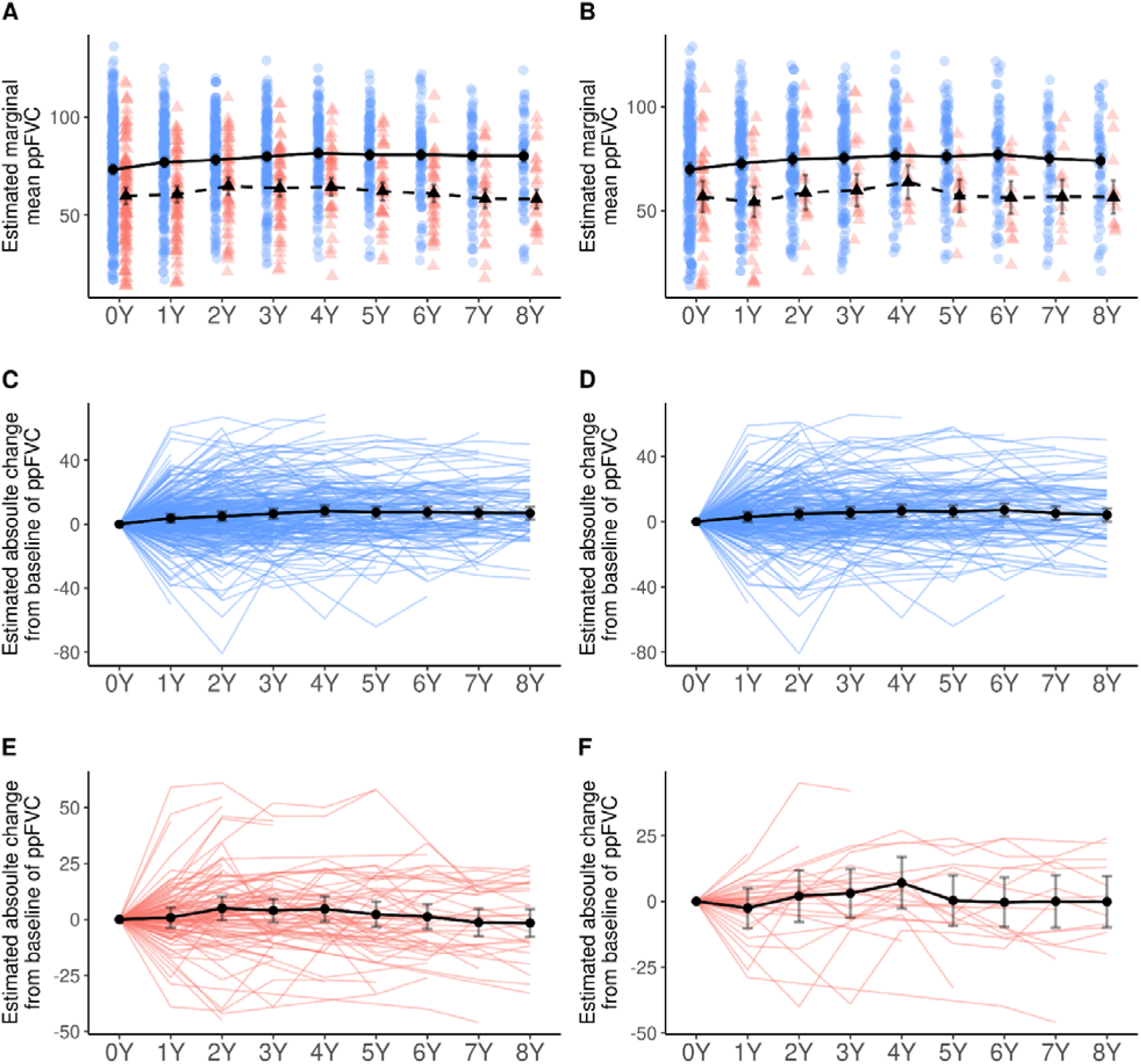
Estimated marginal means (EMMs) of ppFVC trajectories according to fibrosis status. Panels A,C,E show results using the registry definition; panels B,D,F show results using the trial definition. Top panels (A,B) display EMM ppFVC at each yearly visit with 95% CIs (blue circles/solid line = non-fibrosis; red triangles/dashed line = fibrosis). Middle panels (C,D) display EMM change from baseline PFT (0Y) with 95% CIs. Lower panels (E,F) show individual patient trajectories overlaid with group mean changes from baseline PFT (0Y). Counts of ppFVC measurements per visit (0Y–8Y) were: registry non-fibrosis 415, 220, 199, 169, 134, 110, 91, 73, 60; registry fibrosis 135, 82, 65, 59, 47, 41, 41, 25, 27; trial non-fibrosis 291, 170, 160, 141, 110, 93, 78, 59, 59; trial fibrosis 45, 29, 19, 18, 15, 16, 16, 12, 12. Full model-based contrast estimates are provided in Supplemental Tables 2 and 4 (between-group) and 3 and 5 (within-group).

Findings using the trial definition closely mirrored those under the registry definition (Figure 2). At baseline PFT (0Y), the estimated marginal mean ppFVC was 69.9% (95% CI: 66.9,72.9) in the non-fibrosis group and 56.8% (95% CI: 49.2,64.4) in the fibrosis group (mean difference: −13.1%, 95% CI: −21.2, −5.0; p=0.0034). These between-group differences remained statistically significant at all follow-up visits, ranging from −12.7% to −20.6% over 1–8Y (all adjusted p≤0.0039; Supplemental Table 4). Within-group results were also similar. The non-fibrosis group showed increases from baseline PFT (0Y) at several visits (2–6Y: +4.9% to +7.2%; all adjusted p≤0.0075), whereas the fibrosis group showed no significant within-group differences between any visits (Supplemental Table 5).

### Survival

At the end of follow-up 16% of the patients had died or undergone lung transplantation and the likelihood of survival was about 70% (Fig. 3). Applying the register definition of fibrosis, no differences were observed for children with or without fibrosis up to the age of 20 years. (Fig. 3, A). Similar findings were observed for patients having or not having fibrosis according to the trial definition (Fig. 3, B). Comparing the survival of the two cohorts with fibrosis, no differences depending on the fibrosis definition were identified (Fig. 3C).

**Figure 3.**
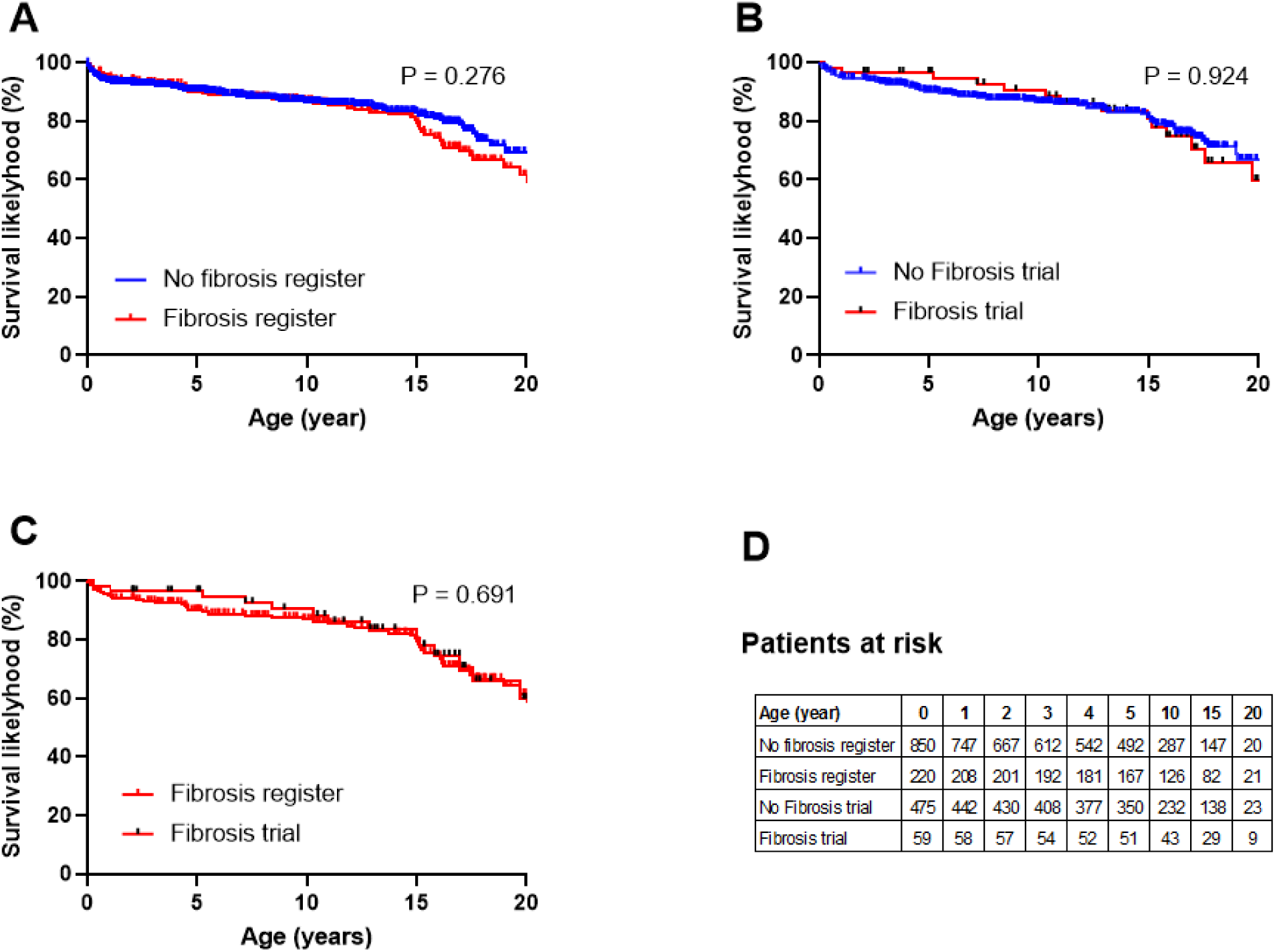
Kaplan-Meier survival curves (age at death or lung transplant) were calculated for the 4 groups and compared by log-rank (Mantel-Cox) test. No differences in survival were observed between patients with or without fibrosis or between patients with the two fibrosis definitions.

## Discussion

This is the first study to assess the prevalence, characteristics and disease trajectories of pulmonary fibrosis in chILD. The studýs strength lies in its systematic application of pulmonary fibrosis definitions to a large group of prospectively followed, well-defined patients representing the entire chILD spectrum. We determined the childhood prevalence of fibrosis to 20.5% using a single time point (register definition) and to 11.6% using a dual time point (trial definition). Among all children able to perform pulmonary function tests, children with fibrosis had 15-20% lower ppFVC than those without, at any time point during childhood. The long-term disease trajectories did not differ between children with or without pulmonary fibrosis throughout childhood. Overall, survival until the age of 20 years was about 70%, with no differences between children with or without fibrosis or between the two fibrosis definitions.

The development of fibrosis may take considerable time and the cumulative contribution of additional triggers including environmental agents or exacerbations ^25^. This is in agreement with our data on pulmonary function testing results. The deterioration in patients with fibrosis likely occurs insidiously and early, as the stable 15-20% lower FVC observed throughout childhood was already established when the first pulmonary function tests were performed. These children were also significantly older and their disease was detected later. The diagnostic gap when routine lung function testing is not yet feasible, highlights the potentially great importance of CT imaging. Beyond the necessity of limiting exposure to radiation, the requirement for general anesthesia to obtain appropriate quality images hinders the feasibility of repeated procedures. Further studies are needed to define standardized imaging protocols and reasonable monitoring intervals. Such studies evaluate the sensitivity of CT compared to FVC measurements. The results of this study indicate that the overall changes of ppFVC suggest a relatively slow progression of impaired lung physiology that is detectable by spirometry. This finding is consistent with the observed average annual ppFVC declines of 1 to 2% in children and early adolescence with ABCA3 deficiency ^26^. These observations emphasize the unresolved challenge of defining progressive pulmonary fibrosis in children, and, determining optimal methods for monitoring disease progression during childhood.

It is expected that the development of fibrosis may also differ with the underlying etiological entity. Previously, analysis of the chILD-EU and US-chILD registries identified 47 different entities in which fibrosis was reported, but without using a uniform definition and not able to give disease specific frequencies ^7^. In the present study, we defined the prevalence of fibrosing lung disease both at the diagnosis and within disease subcategories and identified patients across 72 entities fulfilling criteria for fibrosis. For some entities with sufficient sample size, we estimated prevalence rates above 10%. These included ILD-related to the alveolar surfactant region including ABCA3 and SP-C deficiency and interestingly, ILD secondary to post-infectious processes such as constrictive bronchiolitis, i,e, bronchiolocentric fibrosis ^27^.

To date, there is no comprehensive data on the individual criteria to define pulmonary fibrosis in children. This is particularly relevant for cystic lesions on CT imaging. Cysts are not a criterion in adult fibrotic lung disease, but they are a prominent feature of surfactant dysfunction and many other disorders in childhood, all known to result in pulmonary fibrosis ^7^.

In this study, we additionally assessed the impact of cystic lesions as an indicator of fibrosis. Cystic lesions were the second most frequent CT criterion noted (in 51 to 60%) and have been shown to increase with time in patients with surfactant dysfunction disorders and fibrosis lung disease ^11–14,26^. Including the criterion of cystic lesions as a fibrotic criterion increased the number of fibrosis cases by about 20% (21.8% register definition, 17.7% trial definition). At baseline PFT (0Y), fibrotic patients with a cystic lesion and one another fibrotic lesion on CT had better ppFVC than those with two non-cystic fibrotic lesions on their CT scans, but this difference disappeared over time, suggesting that progression of cystic lesions is associated with accelerated lung function decline. The main diagnostic categories included surfactant dysfunction disorders (n = 19) and cystic lung diseases (n =26), including STING-associated vasculopathy or ARS-deficiencies.

This study applied the currently proposed criteria for defining pulmonary fibrosis in a large prospective multinational cohort of patients with chILD providing rare insight into long-term disease evolution in very rare pediatric lung diseases. All CT scans and lung biopsies submitted to the chILD-EU registry were scored by dedicated peer review teams and use of standardized peer-reviewed imaging and histopathological scoring ensured consistency across centers. Integration of clinical, radiological, and functional data allowed a multidimensional assessment of fibrosis presence and disease progression. Nonetheless, several limitations must be acknowledged. First, referral bias of certain conditions to the registry may have led to under- or overrepresentation of some chILD conditions. Second, the entire cohort of the registry was included in our study, not only those entities typically expected to develop pulmonary fibrosis. While we identified proven pulmonary fibrosis among the less-typical entities, they may represent a distinct type of pulmonary fibrosis with slower progression and a potential reversibility after removal of a noxious exposure. Third, the decision to order CT scans or biopsies was made at the discretion of the treating physician, which could influence detection rates. To overcome this shortage, our comparison group included only children who had the same diagnostic opportunity, i.e. had a biopsy and one CT scan, or more CT scans.

To summarize, this study applied a systematic definition of pulmonary fibrosis and analyzed prospectively collected clinical data in chILD. The study identified disease entities prone to fibrosis and quantified their relative risk. Although survival through childhood did not differ between children with or without fibrosis, those with fibrosis exhibited substantially lower ppFVC, defining them an at-risk group. The consistent application of clear fibrosis criteria will facilitate earlier recognition, better disease classification, and ultimately improved treatment strategies for selected chILD patients.

## Contributors

MG led project administration and planned the study, NS and ES participated in the design of the study. SR-H peer-reviewed lung pathology, JL-Z, BK, IK-S peer-reviewed chest CT imaging, JC, PM, JR, KM-S, CKR, FG, HM, JL, KK, AMR, FB, FS, PSJ, JT, MP, TS, AA, NE, NK, SH, AK, FPr, AW, AM-G, SM, JMB, LN, MW, AM and MK participated in the recruitment of study participants and collected their data. All the chILD-EU collaborators contributed some study participants and their data. JR, KM-S, N-BT and MG performed register quality control, SM, N-BT and FP performed the data analysis, MG, ES and FB prepared the report for publication. All authors have access to all raw data used in this manuscript and had final responsibility for the decision to submit for publication. MG, ES, N-BT and SM have accessed and verified the underlying data.

## Supporting information

Strobe checklist

## Declaration of interest

MG reports personal fees for advisory and adjudication board work from Boehringer Ingelheim, outside the submitted work; grant support from Boehringer to the institution. AM-G reports fees to the institution for work as local principal investigator in a study by Boehringer, AM reports personal fees from Vertex for consulting, JL reports fees to the institution from Boehringer for work as a sub-investigator, HM reports fees to the institution from Boehringer, JLZ reports personal honoraria for lectures and consultation from Boehringer and from Bayer for study evaluation. FP reports personal fees for lectures from Sanofi/Regeneron, Allergopharma, Takeda Pharma, BioCryst, Vertex Pharmaceuticals, Nutricia-Milupa, Stallergenes Greer, ALK-Abelló and for participation in an advisory board from Takeda Pharma and Sanofi/Regeneron. FS reports personal fees for attending meetings and for lectures by Vertex and to the institution by Vertex. FP is founder and partner of RPACT GbR, a company proving statistical analysis for the University of Munich. KK reports personal fees and payments to the institution from Boehringer, NS reports personal fees and payments to the institution from Boehringer.

## Data sharing

Anonymized aggregated data from the chILD-EU register are available upon reasonable request. Research proposals using such aggregated data can be submitted to the corresponding author. The proposals will be assessed by the chILD-EU register coordination team, and aggregated data will be shared if the proposal addresses relevant research questions. Requests for anonymized individual patient data can be sent to the corresponding author, who will facilitate contact with the responsible physicians who will discuss the legal and administrative feasibility of sharing those data.

## Acknowledgments

We thank all parents and children for participating in the study and researchers, technicians, paediatricians who helped with data collection.

## SUPPLEMENTAL MATERIAL

Supplemental statistical methods on longitudinal analysis

Longitudinal analyses of ppFVC were conducted using linear mixed-effects models. Repeated values within the same year were averaged and yearly means were analyzed as categorical time points. The fixed part of the model included fibrosis status (Yes/No), visit, age at baseline PFT (0Y), and the interaction between fibrosis status and visit. A patient-specific random intercept was included to account for within-subject variability. Alternative residual correlation and variance structures were systematically evaluated. We compared first-order autoregressive (AR1), compound symmetry, and fully unstructured forms. Model fit was assessed using maximum likelihood estimation and information criteria (AIC and BIC). The unstructured correlation structure offered flexibility but did not converge reliably given the sparsity of later follow-ups. Compound symmetry converged but showed poorer fit. The autoregressive structure achieved stable convergence and favorable fit and was therefore used. The autoregressive structure corresponds to the assumption that correlations between repeated measures decay exponentially with the temporal lag, which is a plausible pattern for annual lung function follow-up. Residual heteroscedasticity was examined in the same way. Allowing variance to differ by visit improved model fit compared to a homoscedastic specification and was included in the final models. Final models were estimated using restricted maximum likelihood. Missing data were assumed to be missing at random. Estimated marginal means were used to describe within-group trajectories, compare fibrosis groups at each visit, and estimate change from baseline PFT (0Y). Multiple testing was controlled by Tukey adjustment for within-group comparisons across visits and Holm adjustment across visits for the between-group contrasts. Model diagnostics were inspected to check assumptions. These included residual-versus-fitted plots to assess homoscedasticity, quantile–quantile plots for residual normality and distribution of random intercepts. None of these suggested major violations. For sensitivity analyses we repeated the analyses truncating follow-up at 6 years to mitigate sparsity at later visits. Results for betweenLgroup contrasts and withinLgroup changes were materially unchanged.

## SUPPLEMENTAL FIGURE

**Supplemental figure 1.**
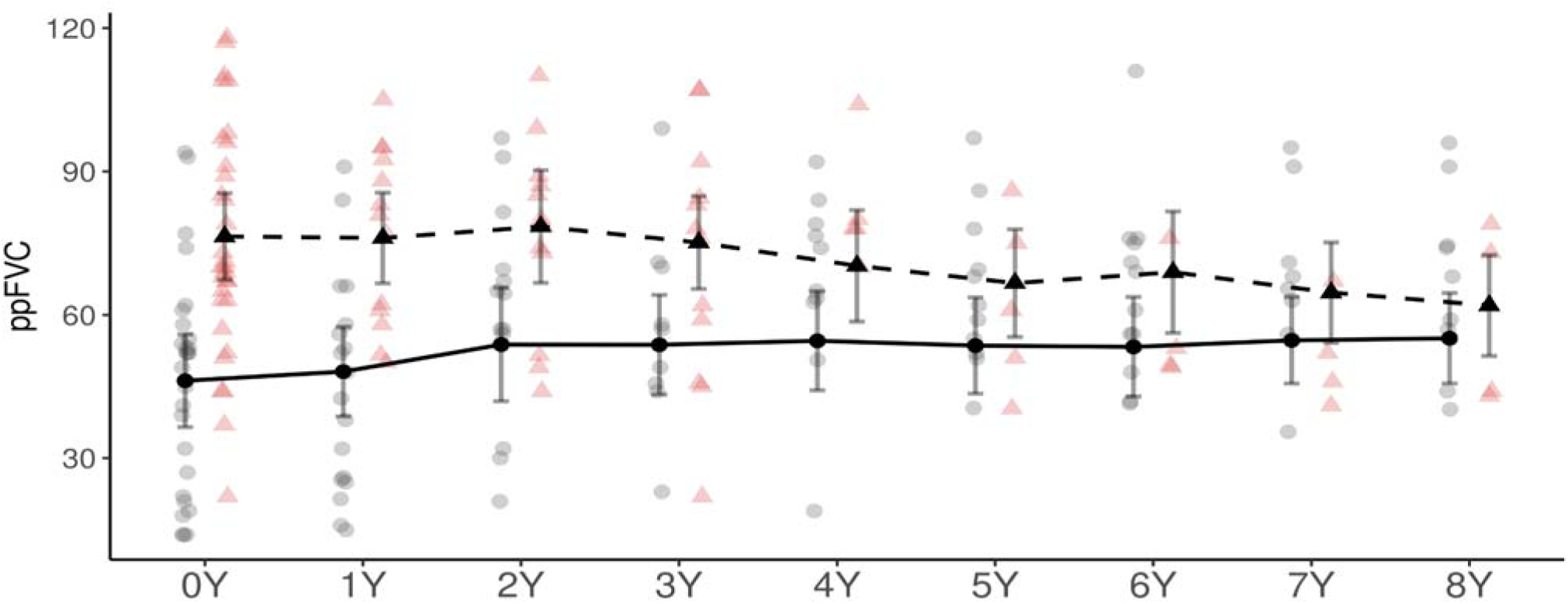
ppFVC over 8 years in patients with cysts and without cysts in CT scans. Estimated marginal means of ppFVC with 95% CIs, obtained from the mixed-effects model, are shown for patients with cysts in CT scans (solid circles, solid line) and without cysts in CT scans (triangles, dashed line) from baseline PFT (0Y) to 8 years of follow-up. Only patients with two fibrosis features on any CT scan were compared. Semi-transparent points represent individual observed values.

## SUPPLEMENTAL TABLES

**Supplemental Table 1.** Country of Origin (N, % total)

|  |  |
| --- | --- |
| Austria | 17 (1.6%) |
| Belgium | 18 (1.7%) |
| Czechia | 5 (0.5%) |
| Denmark | 12 (1.1%) |
| Germany | 602 (56.2%) |
| Greece | 11 (1%) |
| Hungary | 12 (1.1%) |
| Italy | 29 (2.7%) |
| Poland | 86 (8%) |
| Spain | 27 (2.5%) |
| Switzerland | 32 (3%) |
| Turkey | 109 (10.2%) |
| United Kingdom | 94 (8.8%) |
| Other | 17 (1.6%) |
| Total | 1071 (100%) |

**Supplemental Table 2:** Pairwise differences in ppFVC between fibrosis groups using registry definition at all time points. Estimated marginal means for ppFVC were obtained from a linear mixed-effects model adjusted for age at baseline PFT (0Y). Pairwise comparisons between individuals with and without fibrosis were conducted at each time point. P-values were adjusted for multiple testing across time points using the Holm correction. Negative values indicate that individuals with fibrosis had a lower ppFVC compared to those without fibrosis

| Visit | Comparison | Difference in ppFVC (95% CI) | Adjusted p-value |
| --- | --- | --- | --- |
| 0Y | Yes - No | -13.4 (-18.4, -8.5) | <0.0001 |
| 1Y | Yes - No | -16.3 (-21.1, -11.5) | <0.0001 |
| 2Y | Yes - No | -13.3 (-18.5, -8.2) | <0.0001 |
| 3Y | Yes - No | -16.1 (-21.0, -11.1) | <0.0001 |
| 4Y | Yes - No | -17.2 (-22.4, -11.9) | <0.0001 |
| 5Y | Yes - No | -18.7 (-24.0, -13.4) | <0.0001 |
| 6Y | Yes - No | -19.7 (-25.0, -14.4) | <0.0001 |
| 7Y | Yes - No | -21.9 (-27.5, -16.2) | <0.0001 |
| 8Y | Yes - No | -22.0 (-27.8, -16.2) | <0.0001 |

**Supplemental Table 3:** Estimated differences in ppFVC over time within each fibrosis group (Yes/No) using registry definition. Estimated differences in ppFVC between follow-up visits were obtained from a linear mixed-effects model adjusted for age at baseline PFT (0Y). Pairwise comparisons across time points within each fibrosis group were performed using Tukey’s method to adjust for multiple comparisons. Positive values indicate an increase in ppFVC over time, while negative values indicate a decrease.

| Fibrosis by register | Contrast | Difference in ppFVC (95% CI) | P-value |
| --- | --- | --- | --- |
| No | 1Y - 0Y | 3.7 (0.6, 6.8) | 0.0082 |
| No | 2Y - 0Y | 4.9 (1.4, 8.4) | 0.0006 |
| No | 2Y - 1Y | 1.2 (-2.0, 4.4) | 0.9642 |
| No | 3Y - 0Y | 6.6 (3.1, 10.2) | <0.0001 |
| No | 3Y - 1Y | 2.9 (-0.3, 6.2) | 0.1192 |
| No | 3Y - 2Y | 1.7 (-1.4, 4.9) | 0.7417 |
| No | 4Y - 0Y | 8.4 (4.6, 12.2) | <0.0001 |
| No | 4Y - 1Y | 4.7 (1.1, 8.3) | 0.0017 |
| No | 4Y - 2Y | 3.5 (-0.1, 7.1) | 0.0677 |
| No | 4Y - 3Y | 1.8 (-1.4, 4.9) | 0.735 |
| No | 5Y - 0Y | 7.5 (3.5, 11.5) | <0.0001 |
| No | 5Y - 1Y | 3.8 (0.1, 7.5) | 0.0436 |
| No | 5Y - 2Y | 2.6 (-1.2, 6.4) | 0.4585 |
| No | 5Y - 3Y | 0.9 (-2.6, 4.3) | 0.9977 |
| No | 5Y - 4Y | -0.9 (-4.3, 2.5) | 0.996 |
| No | 6Y - 0Y | 7.5 (3.5, 11.6) | <0.0001 |
| No | 6Y - 1Y | 3.8 (0.0, 7.7) | 0.0558 |
| No | 6Y - 2Y | 2.6 (-1.3, 6.6) | 0.498 |
| No | 6Y - 3Y | 0.9 (-2.8, 4.6) | 0.998 |
| No | 6Y - 4Y | -0.9 (-4.6, 2.8) | 0.9985 |
| No | 6Y - 5Y | 0.0 (-3.4, 3.5) | 1 |
| No | 7Y - 0Y | 7.1 (2.9, 11.3) | <0.0001 |
| No | 7Y - 1Y | 3.4 (-0.6, 7.4) | 0.1777 |
| No | 7Y - 2Y | 2.2 (-1.9, 6.3) | 0.7749 |
| No | 7Y - 3Y | 0.4 (-3.4, 4.3) | 1 |
| No | 7Y - 4Y | -1.3 (-5.2, 2.6) | 0.9816 |
| No | 7Y - 5Y | -0.4 (-4.2, 3.4) | 1 |
| No | 7Y - 6Y | -0.5 (-4.1, 3.2) | 1 |
| No | 8Y - 0Y | 6.9 (2.4, 11.5) | 0.0001 |
| No | 8Y - 1Y | 3.2 (-1.2, 7.6) | 0.3507 |
| No | 8Y - 2Y | 2.0 (-2.4, 6.5) | 0.8936 |
| No | 8Y - 3Y | 0.3 (-4.0, 4.6) | 1 |
| No | 8Y - 4Y | -1.5 (-5.8, 2.9) | 0.9818 |
| No | 8Y - 5Y | -0.6 (-4.8, 3.7) | 1 |
| No | 8Y - 6Y | -0.6 (-4.7, 3.6) | 1 |
| No | 8Y - 7Y | -0.1 (-4.1, 3.9) | 1 |
| Yes | 1Y - 0Y | 0.8 (-4.4, 6.0) | 0.9999 |
| Yes | 2Y - 0Y | 5.0 (-1.1, 11.1) | 0.21 |
| Yes | 2Y - 1Y | 4.2 (-1.2, 9.6) | 0.2838 |
| Yes | 3Y - 0Y | 4.0 (-2.0, 10.0) | 0.4942 |
| Yes | 3Y - 1Y | 3.2 (-2.3, 8.6) | 0.6791 |
| Yes | 3Y - 2Y | -1.0 (-6.5, 4.5) | 0.9997 |
| Yes | 4Y - 0Y | 4.7 (-1.8, 11.1) | 0.3745 |
| Yes | 4Y - 1Y | 3.8 (-2.2, 9.9) | 0.5551 |
| Yes | 4Y - 2Y | -0.3 (-6.6, 5.9) | 1 |
| Yes | 4Y - 3Y | 0.7 (-4.7, 6.1) | 1 |
| Yes | 5Y - 0Y | 2.3 (-4.3, 8.8) | 0.9785 |
| Yes | 5Y - 1Y | 1.4 (-4.7, 7.6) | 0.9985 |
| Yes | 5Y - 2Y | -2.8 (-9.2, 3.7) | 0.9237 |
| Yes | 5Y - 3Y | -1.7 (-7.6, 4.2) | 0.9925 |
| Yes | 5Y - 4Y | -2.4 (-7.9, 3.1) | 0.913 |
| Yes | 6Y - 0Y | 1.3 (-5.2, 7.7) | 0.9996 |
| Yes | 6Y - 1Y | 0.4 (-5.6, 6.5) | 1 |
| Yes | 6Y - 2Y | -3.7 (-10.1, 2.6) | 0.6675 |
| Yes | 6Y - 3Y | -2.7 (-8.7, 3.2) | 0.8902 |
| Yes | 6Y - 4Y | -3.4 (-9.3, 2.5) | 0.6891 |
| Yes | 6Y - 5Y | -1.0 (-6.3, 4.3) | 0.9997 |
| Yes | 7Y - 0Y | -1.3 (-8.5, 5.8) | 0.9997 |
| Yes | 7Y - 1Y | -2.2 (-9.0, 4.6) | 0.987 |
| Yes | 7Y - 2Y | -6.4 (-13.4, 0.7) | 0.1211 |
| Yes | 7Y - 3Y | -5.3 (-12.0, 1.3) | 0.2377 |
| Yes | 7Y - 4Y | -6.0 (-12.8, 0.7) | 0.1266 |
| Yes | 7Y - 5Y | -3.6 (-10.1, 2.9) | 0.7256 |
| Yes | 7Y - 6Y | -2.6 (-8.4, 3.2) | 0.9 |
| Yes | 8Y - 0Y | -1.6 (-8.8, 5.6) | 0.999 |
| Yes | 8Y - 1Y | -2.4 (-9.3, 4.5) | 0.9755 |
| Yes | 8Y - 2Y | -6.6 (-13.7, 0.5) | 0.0968 |
| Yes | 8Y - 3Y | -5.6 (-12.3, 1.1) | 0.1972 |
| Yes | 8Y - 4Y | -6.3 (-13.1, 0.6) | 0.1015 |
| Yes | 8Y - 5Y | -3.9 (-10.4, 2.7) | 0.6655 |
| Yes | 8Y - 6Y | -2.9 (-9.2, 3.4) | 0.8928 |
| Yes | 8Y - 7Y | -0.3 (-6.5, 6.0) | 1 |

**Supplemental Table 4:** Pairwise differences in ppFVC between fibrosis groups using trial definition at all time points. Estimated marginal means for ppFVC were obtained from a linear mixed-effects model adjusted for age at baseline PFT (0Y). Pairwise comparisons between individuals with and without fibrosis were conducted at each time point. P-values were adjusted for multiple testing across time points using the Holm correction. Negative values indicate that individuals with fibrosis had a lower ppFVC compared to those without fibrosis

| Visit | Comparison | Difference in ppFVC (95% CI) | Adjusted p-value |
| --- | --- | --- | --- |
| 0Y | Yes - No | -13.1 (-21.2, -5.0) | 0.0034 |
| 1Y | Yes - No | -18.6 (-26.3, -10.8) | <0.0001 |
| 2Y | Yes - No | -15.9 (-24.8, -7.0) | 0.0014 |
| 3Y | Yes - No | -15.6 (-23.8, -7.3) | 0.001 |
| 4Y | Yes - No | -12.7 (-21.4, -4.1) | 0.0039 |
| 5Y | Yes - No | -19.0 (-27.5, -10.5) | 0.0001 |
| 6Y | Yes - No | -20.6 (-29.1, -12.2) | <0.0001 |
| 7Y | Yes - No | -18.3 (-27.1, -9.5) | 0.0003 |
| 8Y | Yes - No | -17.4 (-26.0, -8.8) | 0.0004 |

**Supplemental table 5.**
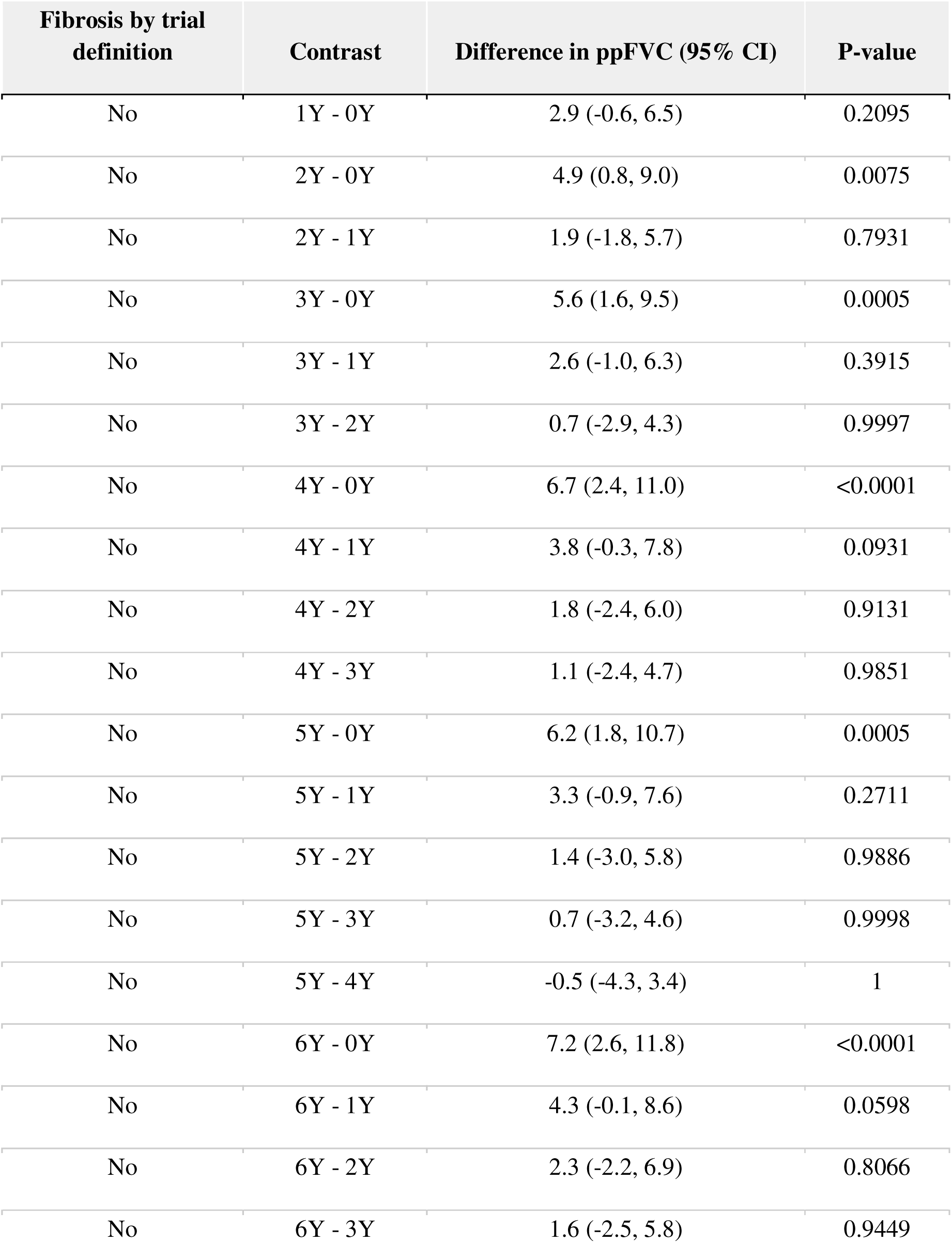
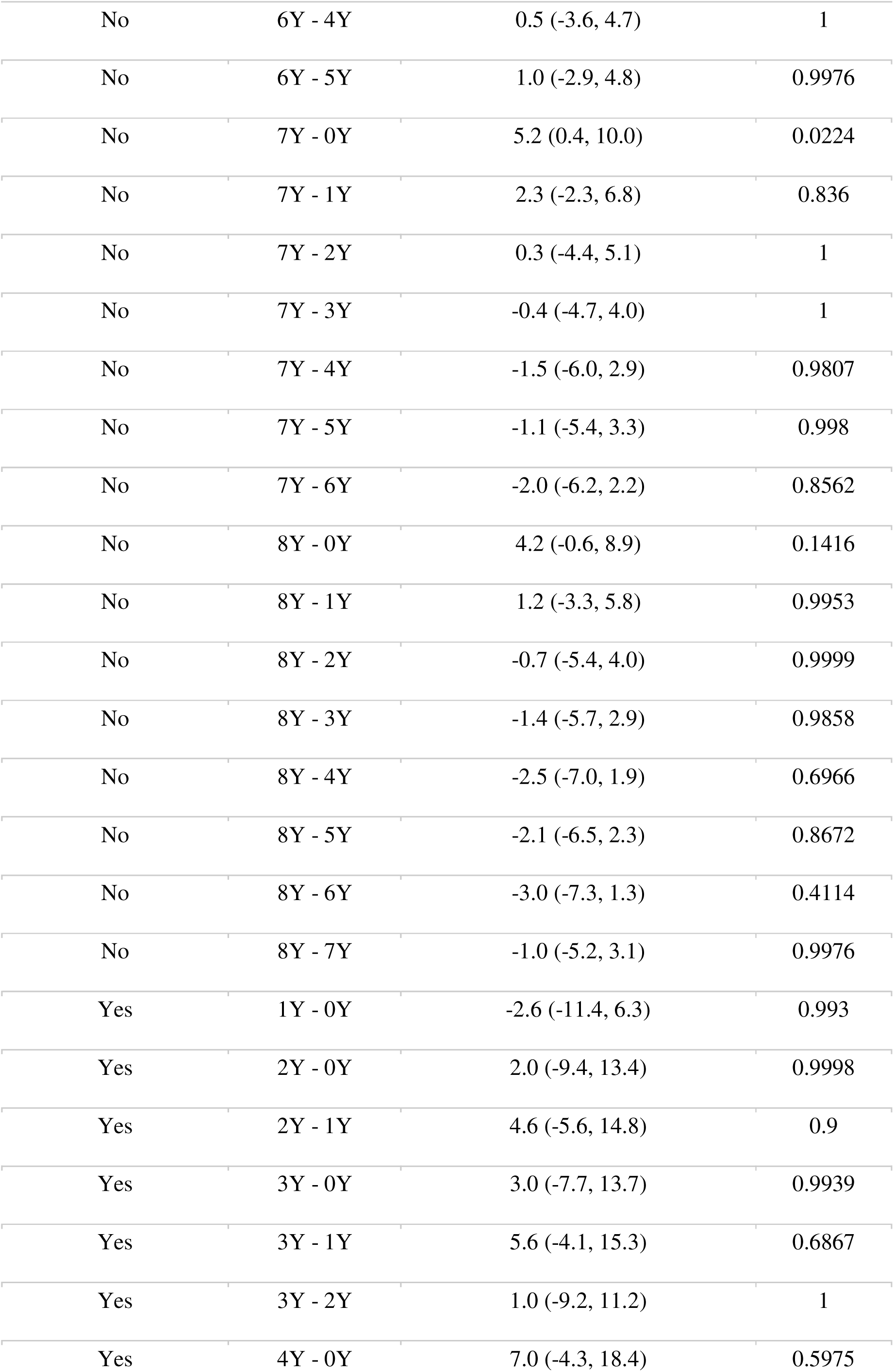
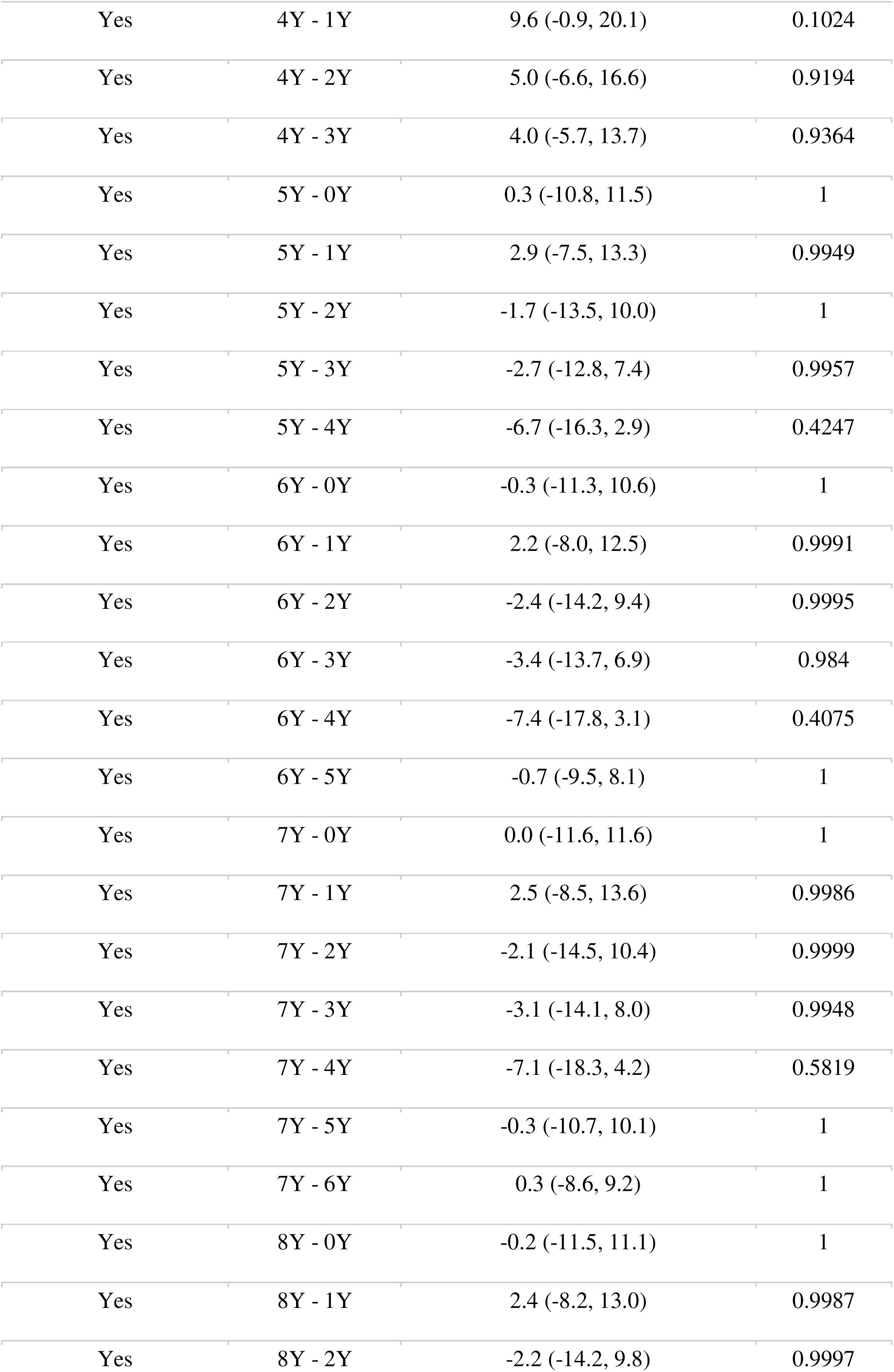
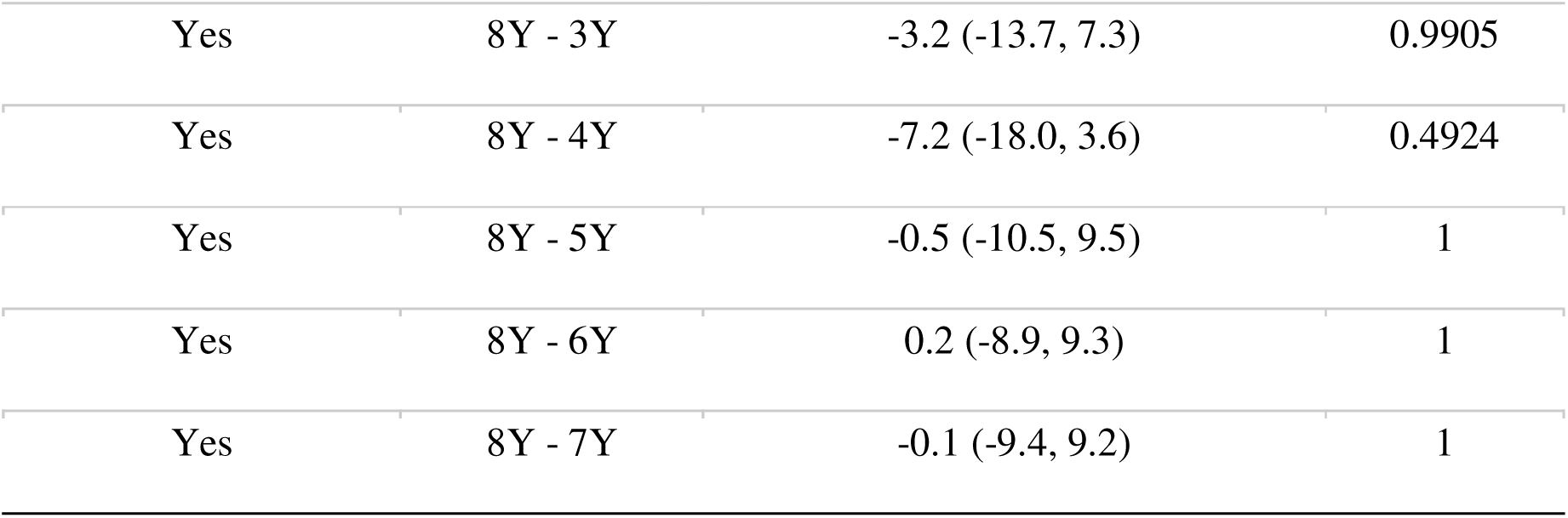
Estimated differences in ppFVC over time within each fibrosis group (Yes/No) using trial definition. Estimated differences in ppFVC between follow-up visits were obtained from a linear mixed-effects model adjusted for age at baseline PFT (0Y). Pairwise comparisons across time points within each fibrosis group were performed using Tukey’s method to adjust for multiple comparisons. Positive values indicate an increase in ppFVC over time, while negative values indicate a decrease.

## Notes

### Author Declarations

Participating centers adhered to all contractual, legal, and ethical standards before enrolling patients. Each patient and/or caregiver provided age-appropriate verbal assent and written informed consent. The study was approved by the lead Ethics committee of the University Hospital Munich (20-0329, 22-0133) and all local committees.

