## Supplementary material for "Fibrosing interstitial lung disease in childhood: prevalence and disease trajectories": Supplmentary figures

Supplemental figure 1

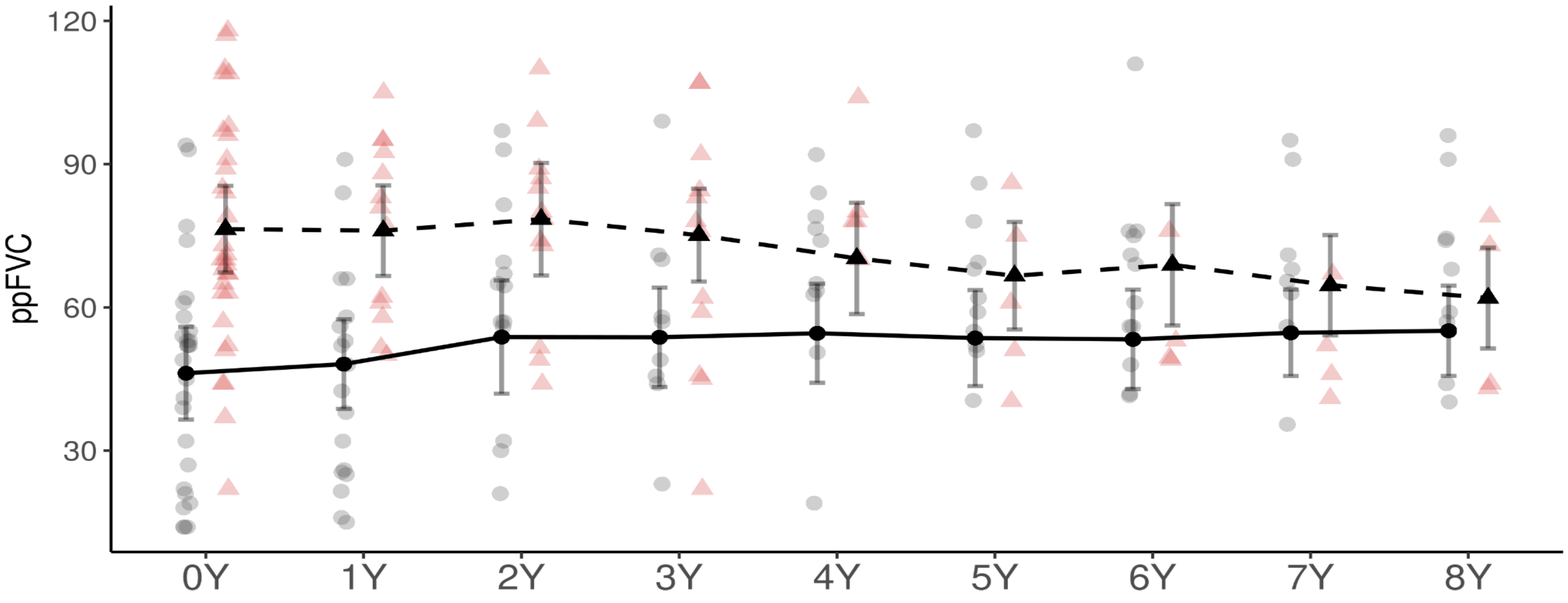


**Supplemental figure 1. ppFVC over 8 years in patients with cysts and without cysts in CT scans.** Estimated marginal means of ppFVC with 95% CIs, obtained from the mixed-effects model, are shown for patients with cysts in CT scans (solid circles, solid line) and without cysts in CT scans (triangles, dashed line) from baseline (0 years) to 8 years of follow-up. Only patients with two fibrosis features on any CT scan were compared. Semi-transparent points represent individual observed values.
